# Mitochondrial genome variants associated with Amyotrophic Lateral Sclerosis and their haplogroup distribution

**DOI:** 10.1101/2024.04.23.24306232

**Authors:** Marcelo R. S. Briones, João H. Campos, Renata C. Ferreira, Lisa Schneper, Ilda M. Santos, Fernando M. Antoneli, NYGC ALS Consortium, James R. Broach

**Affiliations:** Center for Medical Bioinformatics, Escola Paulista de Medicina, Federal University of São Paulo, São Paulo, SP, Brazil, CEP 04039032; Graduate Program in Microbiology and Immunology, Federal University of São Paulo, São Paulo, SP, Brazil, CEP 04039032; Department of Neurology and Neurosurgery, Federal University of São Paulo, São Paulo, SP, Brazil, CEP 04039032; Bridges Genomics, M.E., São Paulo, SP, Brazil, CEP 043639040; Institute for Personalized Medicine, Department of Biochemistry, Pennsylvania State University College of Medicine, 500 University Drive, 17033, Hershey PA, USA; The New York Genome Center (NYGC) ALS Consortium members are listed in the Supplemental Material

**Keywords:** Amyotrophic Lateral Sclerosis, mitochondrial genome, SNVs, GWAS, mitochondrial haplogroups

## Abstract

**Introduction/Aims:** Amyotrophic lateral sclerosis (ALS) may be familial or sporadic, and twin studies have revealed that even sporadic forms have a significant genetic component. Variants in 55 nuclear genes have been associated with ALS and although mitochondrial disfunction is observed in ALS, variants in mitochondrial genomes (mitogenomes) have not yet been tested for association with ALS.

**Methods:** Here we conducted a genome wide association study (GWAS) in mitogenomes of 1,965 ALS patients and 2,547 controls to test the hypothesis that mitogenome variants are associated with ALS.

**Results:** We identified 51 mitogenome variants with *p*-values <10^−7^of which 13 variants have odds ratios (OR)>1, in genes *RNR1*, *ND1*, *CO1*, *CO3*, *ND5*, *ND6* and *CYB*, while 38 variants have OR<1 in genes *RNR1*, *RNA2*, *ND1*, *ND2*, *CO2*, *ATP8*, *ATP6*, *CO3*, *ND3*, *ND4*, *ND5*, *ND6* and *CYB*. The frequencies of haplogroups H, U and L, the most frequent in our ALS dataset, are the same in different onset sites (bulbar, limb, spinal and axial). Also, intra-haplogroup GWAS revealed unique ALS-associated variants in haplogroups L and U.

**Discussion:** Our study shows that mitogenome variants (SNVs) are associated with ALS and suggests that these SNVs could be included in routine genetic testing for ALS and that mitochondrial replacement therapy would have a potential basis for ALS treatment.

## 1 INTRODUCTION

Pathogenic variants in more than 30 nuclear genes have been identified in familial ALS (10% of all ALS patients) ^1^. Although the remaining 90% of patients are apparently sporadic, pathogenic variants identified in familial ALS have also been described in many sporadic patients ^2^. Studies with twins suggest that many patients with sporadic ALS (SALS) may also have an underlying genetic cause ^3^. Although ALS heritability is 40-60%, with 5-10% family history, the typical Mendelian inheritance pattern is not observed ^4^. Among the principal causes of non-Mendelian, or extrachromosomal, inheritance, mitochondrial DNA is a major player, especially in muscular and neuronal diseases ^5^. ALS is associated with abnormal morphology and bioenergetics of mitochondria, which contribute to the denervation of muscles in early stages of ALS ^6^. The mitochondrial respiratory chain contains components expressed by two genomes, nuclear and mitochondrial, and therefore mutations in both genomes affect cellular respiration ^7^. The mitochondrial genome has a mutation rate that is 10 to 20-fold higher than the nuclear genome ^8^. To date, over 250 pathogenic mtDNA mutations (point mutations and rearrangements) have been described ^9^. As many as 1 in 10,000 people have a clinically manifested mtDNA disease and 1 in 6,000 are at risk ^7^. Relevant diseases with mtDNA mutations include dystonia, encephalomyopathy, Alzheimer’s disease and Parkinson’s disease, which are associated with mutations in mitochondrial encoded complex I NADH dehydrogenase subunits ^10^.

All genes associated with ALS so far identified, are nuclear encoded and although mitochondrial dysfunction is observed in ALS, no ALS-associated mutations in mitochondrial genomes have been identified. A mutation in a nuclear gene encoding a mitochondrial protein (*CHCHD10*) is linked to ALS-FTD (frontotemporal dementia) ^11^. An initial study of mitochondrial DNA sequencing in thirty-eight ALS patients and 42 unaffected controls indicated that ALS patients have a higher mean number of variants in protein-coding genes, such as *ND4L*, *ND5*, *ND6*, and *ATP8* and that mitochondrial haplogroups Y and M7c may modulate the clinical expression of ALS ^12^. We performed a mitochondrial genome case/control study to test if variants in the mitochondrial genome are associated with ALS.

## 2 METHODS

### 2.1 Source of data

ALS samples from Pennsylvania State University College of Medicine were collected and analyzed under IRB protocol. The NYGC ALS data was determined by the IRB on 7/23/2015 not to be human subjects research under The Penn State Personalized Research for Innovation, Discovery, and Education (PRIDE) Program (IRB Protocol No. 40532). The Biomedical Research Alliance of New York (BRANY) Institutional Review Board serves as the central ethics oversight body for the New York Genome Center (NYGC) Amyotrophic Lateral Sclerosis (ALS) Consortium. Ethical approval was given and is effective. The period of time over which the samples were collected is not available. Informed consent forms for all study participants were obtained according to IRB protocols at Penn State and NYGC. Diagnosis, sex, age, disease duration, ethnicity and region of onset of patients here tested are shown in Tables S1 and S2. Description of non-ALS (control) samples are in the 1000 Genomes Project (1KGP) (https://www.internationalgenome.org/) and Wei et al. control dataset ^13^.

A total of 4,512 mitochondrial genomes were included in the main analysis. This dataset included 1,965 mitogenomes from ALS patients diagnosed by investigators at NYCG ALS Consortium participating centers. El Escorial criteria were available for a subset of patients. Sequences of 1,930 patients were obtained by the NYGC ALS Consortium and 35 sequenced in the Institute for Personalized Medicine at the Pennsylvania State University with REPLI-g Mitochondrial DNA Kit (Qiagen), library construction with Kapa Hyper Prep kit and adapters (BioO scientific) and sequencing with Illumina MiSeq 2×300. These sequences have been deposited in GenBank with accession numbers MZ458603-MZ460580. Mitochondrial reads, from samples sequenced at the NYGC, were extracted from whole genome BAM files, using samtools view (v. 1.6.0). The control, non-ALS set, comprised 2,534 mitogenomes from the 1KGP ^14^, phase 3, plus 13 control mitogenomes sequenced in the Institute for Personalized Medicine at the Pennsylvania State University with the same method as for ALS samples (Illumina MiSeq 2×300), totaling 2,547 controls (Table S3). The NYCG samples were prepared using either the Illumina truseq nano library prep or the Illumina PCR free library prep protocol and all sequence reads were obtained using the Illumina HiSeq X-10 platform (Table S4).

### 2.2 Genome assembly

The VCF file containing the samples and SNVs from the 1KGP phase 3, which was generated by the project team using the Python script ‘callMom.v.0.2.py’ (https://github.com/juansearch/callMom/blob/master/callMom.v.0.2.py), was obtained from the project’s official portal: https://ftp.1000genomes.ebi.ac.uk/vol1/ftp/release/20130502/README_chrMT_phase3_callmom.md.

Samples sequenced at NYGC and Penn State were aligned using the NCBI reference sequence for the human mitochondrial genome Revised Cambridge Reference Sequence (rCRS), haplogroup H2a2a1 (NC_012920.1) ^15^, which is the same reference used by the 1KGP. Sequence reads were aligned with BWA-MEM (BWA version 0.7.17 ^16^) with mean coverage 8,000x for NYGC assemblies and 180x for Penn State samples. After initial mapping, BAM files were processed according to GATK best practices: (a) reads were sorted (SortSam), (b) duplicates were marked (MarkDuplicates), and (c) mitochondrial variants were called (HaplotypeCaller, GenomicsDBImport, GenotypeGVCFs) using GATK version 4.0.1.0. This process generated a VCF file with 1,965 mitogenomes from ALS patients and 13 control mitogenomes. To merge this VCF with that of the 1KGP (2,534 mitogenomes), the simple shell script with ‘sed’ command was used to ensure that all samples and SNVs had names consistent with the reference FASTA file for the mitochondrial genome and the mitochondrial contig. After ensuring consistency, the merge of the VCF files was performed using BCFTools in SamTools. Subsequently, variant frequencies of the final cohort were recalculated using PLINK version 1.90b6.4. Admixture percentages were provided by the ALS NYGC Consortium. Figures were generated using R (version 4.3.1) (https://www.r-project.org/about.html), ggplot2 (version 3.5.0) ^17^ and Cairo (version 1.6-2) ^18^. For admixture plot focusing on ALS subjects, subjects assigned to “atypical for GM1 ab”, “control”, “other motor neuron disease or neurological disorders” or “unknown” subject groups in the metadata provided by the consortium were removed.

### 2.3 GWAS statistical analysis

For GWAS statistical analysis, a VCF file was created with case and control sequences. After initial mapping, the resulting BAM files were processed using the GATK best practices quality control ^19^ as follows: (a) all reads were sorted using SortSam algorithm, (b) duplicate reads were marked using MarkDuplicates algorithm (c) mitochondrial variants were called using the HaplotypeCaller, GenomicsDBImport and GenotypeGVCFs algorithms, as implemented in GATK version 4.0.1.0 ^20^. The VCF file generated by this procedure comprised the 1,965 mitogenomes from ALS patients plus the 13 control mitogenomes. This VCF file was merged with the VCF file containing the 2,534 mitogenomes from the 1000 Genomes Project phase 3, using BCFTools package in SamTools ^21,22^. The final merged VCF totaled 4,512 mitogenomes. After filtering by the standard quality control procedure as described by Li and colleagues ^21,22^, a total of 1,946 variants remained.

The statistical analysis was performed using PLINK version 1.90b6.4 ^23^. SNVs with a genotype failure rate >1% were removed. To keep only the common and the low frequency variants we also removed SNVs with Minimum Allele Frequency (MAF) <0.1%. The sex information was not used. A Principal Component Analysis (PCA) was computed to correct for population stratification ^24^. The association study was performed by logistic regression with the first 20 PCA eigenvectors as covariates. Bonferroni correction for multiple testing with the strict threshold of 10^−7^ was employed resulting in 51 variants. Among these, 43 were common variants (MAF ≥1%) and 8 were low frequency variants (MAF >0.1% and <1%). The *p-*values and Odds Ratios were calculated using PLINK. Estimation of Odds Ratios in 2×2 contingency tables with zeros was performed with Haldane-Anscombe correction ^25^. Haplogroup assignment was done using Haplogrep2 ^26^. Variants were annotated using the NCBI Variation Viewer (https://www.ncbi.nlm.nih.gov/variation/view/). Genomic, clinical and functional annotations of all nucleotide changes that cause non-synonymous substitutions in human mitochondrial genes associated with ALS were annotated using MitImpact (https://mitimpact.css-mendel.it/). The Relative Risk was calculated from the Odds Ratios values according to the formula proposed by Zhang and collaborators ^27^

The mitochondrial genomes of 1,978 samples sequenced at the New York Genome Center (1,965 cases and 13 controls) were scanned to detect homoplasmic or heteroplasmic sites using the Mutserve tool (version 2.0.0-rc12) ^28^.

### 2.4 Methods of alternative dataset analysis

In addition to the dataset described in the main text, two additional datasets were used in parallel analyses: 1. From the original dataset of 4,512 individuals, we randomly removed 860 affected individuals from haplogroups with overrepresented cases (H, J, K, T, U), to balance the haplogroup distribution, leaving a total of 3,652 individuals (1,106 cases and 2,546 controls). The same filters of the original analysis were used (removal of SNVs with genotype failure rate >1% and MAF <0.1%) leaving a total of 1,934 variants; 2. We aligned 30,502 mitogenomes from GenBank as described in Wei et al. ^13^. After removal of partial sequences, low quality samples (with Ns) and large gaps in 5’ and 3’ ends, an alignment was obtained for a subset of 17,725 mitogenomes. A VCF file from this multiple alignment was generated and merged with the VCF file containing the 1,979 samples sequenced at the Pennsylvania State University Institute of Personalized Medicine, resulting in a total of 19,701 mitogenomes (1,965 cases and 17,736 controls). Using the same filters as described in the Methods section (removal of SNVs with genotype failure rate >1% and MAF <0.1%), a total of 1,012 variants were obtained. The remaining statistical analysis of these two additional sets followed the same steps of the original dataset of 4,512 individuals.

## 3 RESULTS

### 3.1 Principal Component Analysis

The Principal Component Analysis (PCA) of the 1,965 ALS mitochondrial genomes reveal the expected populational structure with haplogroup L (present in higher frequency in populations with African background) showing significantly increased diversity as compared to haplogroups associated with European and Asian populations (Figure 1). Figures 1B and 1C zoom in, with more detail, the “European/Asian” (e.g., H, K, J, U, M) haplogroups clustering far from L haplogroups in the same PCA. The PCA profile is maintained if the control samples (2,547) are added to the analysis (Figure S1), and the distribution of cases and controls is shown to be quite similar (Figure S2).

**FIGURE 1.**
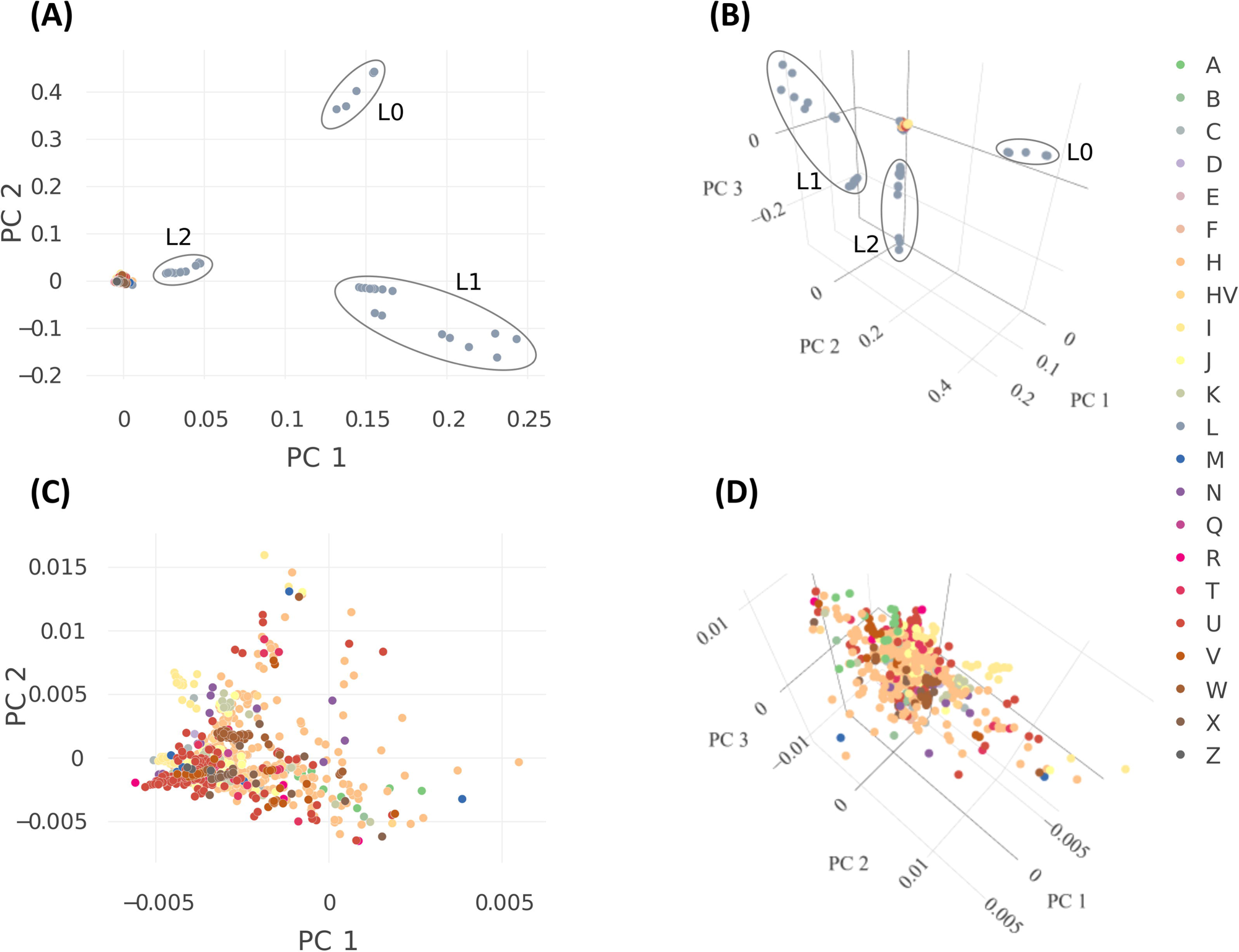
Principal Component Analysis (PCA) of 1,965 ALS cases in this study. The color scale indicates the mitochondrial haplogroups. (A) the 2D PCA, (B) the corresponding 3D PCA with a third component, (C) a closer look at the non-African cluster and (D) the detailed 3D zooms in of the European-Asian haplogroups in the ALS samples. African (L) and Eurasian (EA) haplogroups are highlighted (ovals) in A and B.

The ALS mitogenomes and control genomes were aligned, and variants with a genotyping rate >0.1 or a Minimum Allele Frequency (MAF) <0.1% were removed resulting in the retention of 1,946 variants (SNVs). The QQ-plot of all 1,946 variants show the expected slope increase as compared to the theoretical expected distribution (Figure 2A).

**FIGURE 2.**
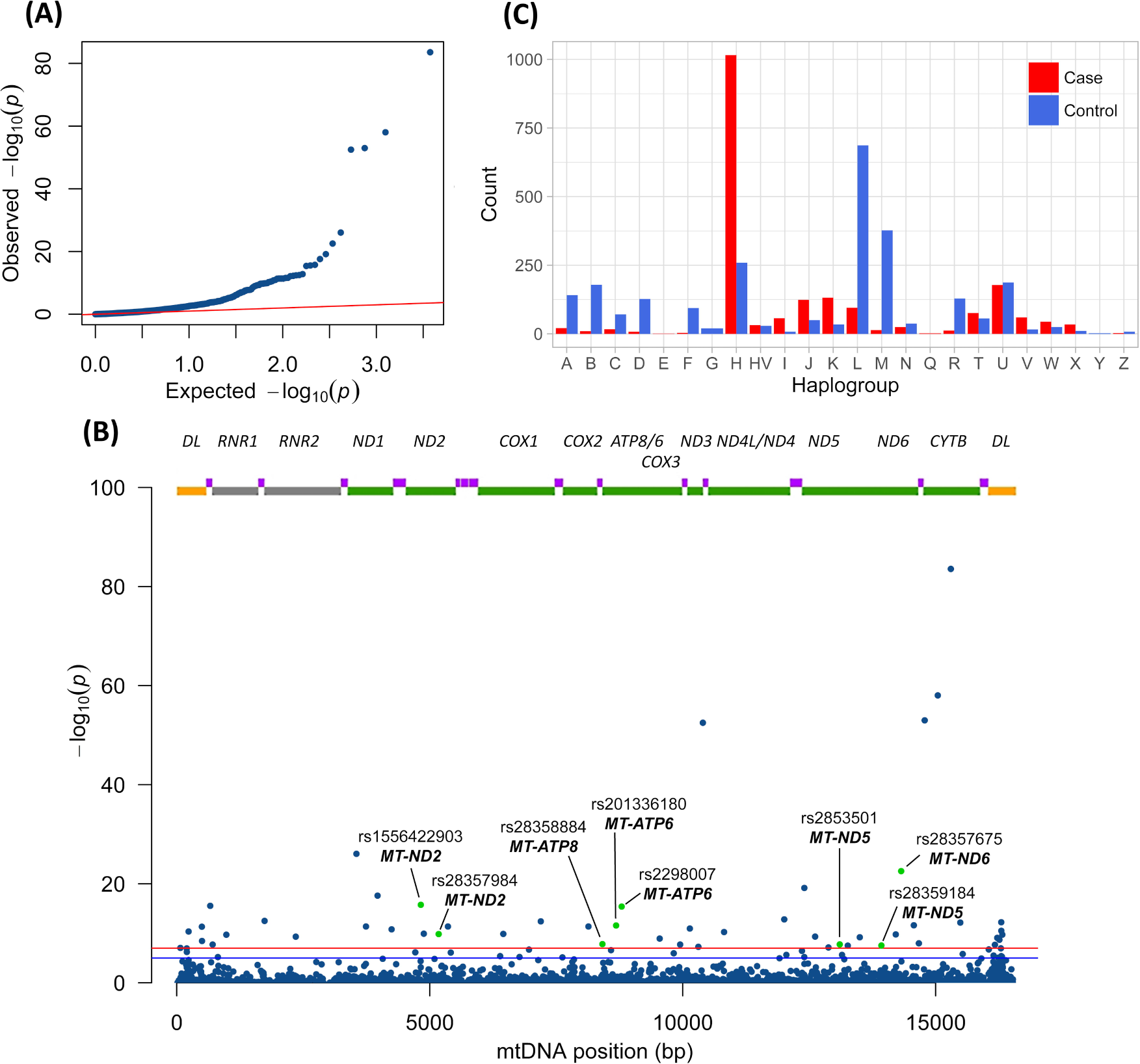
The QQ-Plot (A) with logistic regression of the 1,946 SNVs selected after removal of SNVs with genotype failure rate >0.1 and Minimum Allele Frequency (MAF) <0.1%. The red line is the expected random association values. This corresponds to the analysis of 4,512 samples, being 1,965 cases and 2,547 controls. In (B) Manhattan plot of the 1,946 SNVs mapped to the mitochondrial genome. In the abscissa are the mitochondrial positions numbered according to the revised Cambridge Reference Sequence (rCRS) ^15^ and in the ordinate the −log_10_(*p-*values). The horizontal blue line indicates the 10^−5^ threshold and the red horizontal line the 10^−7^ threshold. The gene map is depicted at the top with yellow boxes representing the D-loop, green boxes the protein coding genes, gray boxes the rRNA genes and purple boxes the tRNA genes. Nonsynonymous substitutions are depicted as green dots and annotated. In (C) Plot of the mitochondrial haplogroup frequencies in 1,965 cases and 2,547 controls. The haplogroups are depicted in the abscissa and the number of samples in each category are depicted in the ordinate.

### 3.2 Distribution of SNVs in the mitogenome

The Manhattan Plot of the 1,946 SNVs shows the distribution of the negative logarithm of the *p*-values of the case-control logistic regression for the corresponding SNVs along the mitogenome. There are 51 variants above the threshold value −log_10_(*p*) = 7 (below the *p-*value 10^−7^) (Figure 2B). These SNVs are significantly associated with ALS and their annotation reveals that they are scattered among almost all mitochondrial genes. For the *p*-value threshold of 10^−5^ there are 73 SNVSs above the threshold, for 5×10^−8^, there are 48 SNVs above the threshold and for 10^−8^, there are 40 SNVs above the threshold. We used the threshold of 10^−7^ because it is 1000-fold below the number of independent tests (10^4^) equivalent to the mitochondrial genome size. Nevertheless, with the threshold of 10^−7^ we are still at a 99.9% significance level per SNV.

### 3.3 Odds Ratios of mitogenome variants

Among these 51 ALS variants, the Odds Ratios (OR) of 13 SNVs are >1 (Table 1) and 38 SNVs show OR <1 (Table 2). The lower bound of confidence intervals of OR >1 are always >1 and in SNVs with OR <1 the upper bound is always <1 indicating that if the study was repeated and the range calculated each time, the true value would lie within these ranges in 95% of occasions. These higher confidence levels ascertain that the interval contains the true odds ratio and that in none of these ORs the confidence interval crosses the threshold value OR =1. The odds ratios in Tables 1 and 2 are corrected for population structure by logistic regression using 20 components from the PCA with cases and controls (Figures S1 and S3). The change in magnitude and direction (from protective to risk increasing), is a direct function of the weights of the components used in the logistic regression. The Relative Risk values, calculated as described by Zhang and colleagues ^27^ to correct the adjusted odds ratios from logistic regression analyses, are included in Tables 1 and 2 for SNVs associated with ALS. The Chi-square tests and Fisher exact test values for Odds Ratios are shown in Table S5.

**TABLE 1.** Mitochondrial genome variants associated with increased ALS risk. Only variants with *p*-value <10^−7^ are shown. Logistic regression of 2,547 controls and 1,965 ALS cases. *MT* = mitochondrial, *HV* = hyper variable region, *CO* = Cytochrome *c* oxidases, *CYB* = Cytochrome *b*, *ND* = NADH ubiquinone oxidoreductases, *RNR1* = 12S ribosomal RNA. Nucleotide changes as shown in PLINK program. Variants annotated using NCBI Variation Viewer. (https://www.ncbi.nlm.nih.gov/variation/view/). 95% CI = 95% confidence interval. Risk = Relative Risk Ratio calculated as proposed by Zhang and collaborators ^27^.

| dbSNP ID | Position | Change | Gene | P-value | Odds Ratio | 95% C.I. | Risk | 95% CI |
| --- | --- | --- | --- | --- | --- | --- | --- | --- |
| rs879158303 | 72 | T>C | MT-HV2 | 8.888x10<sup>-8</sup> | 3.842 | 2.346 - 6.292 | 3.75 | 2.32 - 6.02 |
| rs28660704 | 497 | C>T | MT-HV3 | 4.783x10<sup>-12</sup> | 5.031 | 3.182 - 7.955 | 4.86 | 3.12 - 7.49 |
| rs28358568 | 710 | T>C | MT-RNR1 | 1.897x10<sup>-8</sup> | 361.2 | 46.33 - 2815 | 281.56 | 44.73 - 877.06 |
| rs28358587 | 3552 | T>A | MT-ND1 | 9.02x10<sup>-27</sup> | 1.992x10<sup>4</sup> | 3.255x10<sup>3</sup> - 1.219x10<sup>5</sup> | 35.80 | 35.48 - 35.85 |
| rs28516468 | 6455 | C>T | MT-CO1 | 1.363x10<sup>-10</sup> | 63.44 | 17.87 - 225.2 | 22.15 | 11.88 - 29.28 |
| rs28358875 | 7196 | C>A | MT-CO1 | 3.895x10<sup>-13</sup> | 136.5 | 36.19 - 514.8 | 24.00 | 16.32 - 27.42 |
| rs878853022 | 9545 | A>G | MT-CO3 | 1.201x10<sup>-9</sup> | 39.24 | 12.02 - 128.1 | 17.35 | 8.81 - 24.67 |
| rs1556424102 | 12405 | C>T | MT-ND5 | 7.097x10<sup>-20</sup> | 3.322x10<sup>6</sup> | 1.321x10<sup>5</sup> - 8.354x10<sup>7</sup> | 45.47 | 45.46 - 45.475 |
| rs28359175 | 13263 | A>G | MT-ND5 | 3.154x10<sup>-8</sup> | 111.6 | 21.01 - 593.2 | 25.44 | 13.02 - 31 |
| rs28357675 | 14318 | T>C | MT-ND6 | 2.886x10<sup>-23</sup> | 8969 | 1490 - 5.4x10<sup>4</sup> | 35.72 | 35.04 - 35.84 |
| rs28357370 | 15487 | A>T | MT-CYB | 6.983x10<sup>-13</sup> | 129.3 | 34.28 - 487.4 | 23.75 | 15.92 - 27.31 |
| rs386829303 | 16297 | T>C | MT-HV1 | 5.838x10<sup>-13</sup> | 3.994x10<sup>5</sup> | 1.195x10<sup>4</sup> - 1.334x10<sup>7</sup> | 47.09 | 46.92 - 47.1 |
| rs148377232 | 16298 | T>C | MT-HV1 | 3.139x10<sup>-11</sup> | 4.939 | 3.082 - 7.914 | 4.18 | 2.81 - 6.01 |

**TABLE 2.** Mitochondrial genome variants associated with reduced ALS risk. Only variants with *p*-value < 10^−7^ are shown. Logistic regression of 2,547 controls and 1,965 ALS cases. *MT* = mitochondrial, *HV* = hyper variable region, *CO* = Cytochrome *c* oxidases, *CYB* = Cytochrome *b*, *ND* = NADH ubiquinone oxidoreductases, *ATP6* = mitochondrially encoded ATP synthase membrane subunit 6, *ATP8* = mitochondrially encoded ATP synthase membrane subunit 8, *RNR1* = 12S ribosomal RNA and *RNR2* = 16S ribosomal RNA. Nucleotide changes as shown in PLINK program. Variants annotated using NCBI Variation Viewer. (https://www.ncbi.nlm.nih.gov/variation/view/). CI = confidence interval. Risk = Relative Risk Ratio calculated as proposed by Zhang and collaborators ^27^.

| dbSNP ID | Position | Change | Gene | P-value | Odds Ratio | OR 95% CI | Risk | RR 95% CI |
| --- | --- | --- | --- | --- | --- | --- | --- | --- |
| rs3937037 | 235 | A>G | MT-HV2 | 4.447x10<sup>-11</sup> | 1.7x10<sup>-3</sup> | 2.55x10<sup>-4</sup> - 1.133x10<sup>-2</sup> | 0.00 | 0 - 0.012 |
| rs3901846 | 499 | G>A | MT-HV3 | 3.575x10<sup>-9</sup> | 1.837x10<sup>-2</sup> | 4.837x10<sup>-3</sup> - 6.927x10<sup>-2</sup> | 0.02 | 0.005 - 0.073 |
| rs56489998 | 663 | A>G | MT-RNR1 | 2.879x10<sup>-16</sup> | 2.118x10<sup>-4</sup> | 2.789x10<sup>-5</sup> - 1.609x10<sup>-3</sup> | 0.00 | 0 - 0.002 |
| rs397515731 | 980 | T>C | MT-RNR1 | 1.969x10<sup>-10</sup> | 5.584x10<sup>-2</sup> | 2.296x10<sup>-2</sup> - 1.358x10<sup>-1</sup> | 0.06 | 0.023 - 0.137 |
| rs193303006 | 1736 | A>G | MT-RNR2 | 3.238x10<sup>-13</sup> | 6.873x10<sup>-4</sup> | 9.684x10<sup>-5</sup> - 4.877x10<sup>-3</sup> | 0.00 | 0 - 0.005 |
| rs28358579 | 2352 | T>C | MT-RNR2 | 4.92x10<sup>-10</sup> | 1.462x10<sup>-1</sup> | 7.98x10<sup>-2</sup> - 2.68x10<sup>-1</sup> | 0.16 | 0.086 - 0.283 |
| rs878907222 | 3741 | C>T | MT-ND1 | 4.278x10<sup>-12</sup> | 2.942x10<sup>-2</sup> | 1.085x10<sup>-2</sup> - 7.977x10<sup>-2</sup> | 0.03 | 0.011 - 0.08 |
| rs9629042 | 3970 | C>T | MT-ND1 | 2.551x10<sup>-18</sup> | 1.848x10<sup>-6</sup> | 9.536x10<sup>-8</sup> - 3.58x10<sup>-5</sup> | 0.00 | 0 |
| rs9326618 | 4248 | T>C | MT-ND1 | 1.595x10<sup>-11</sup> | 1.388x10<sup>-3</sup> | 2.048x10<sup>-4</sup> - 9.408x10<sup>-3</sup> | 0.00 | 0 - 0.01 |
| rs15564229903 | 4824 | A>G | MT-ND2 | 1.791x10<sup>-16</sup> | 1.961x10<sup>-4</sup> | 2.571x10<sup>-5</sup> - 1.496x10<sup>-3</sup> | 0.00 | 0 - 0.002 |
| rs200763872 | 4883 | C>T | MT-ND2 | 1.233x10<sup>-10</sup> | 8.86x10<sup>-2</sup> | 4.235x10<sup>-2</sup> - 1.854x10<sup>-1</sup> | 0.09 | 0.044 - 0.193 |
| rs28357984 | 5178 | C>A | MT-ND2 | 1.46x10<sup>-10</sup> | 7.716x10<sup>-2</sup> | 3.525x10<sup>-2</sup> - 1.689x10<sup>-1</sup> | 0.08 | 0.037 - 0.176 |
| rs879217723 | 5360 | C>T | MT-ND2 | 4.278x10<sup>-12</sup> | 2.942x10<sup>-2</sup> | 1.085x10<sup>-2</sup> - 7.977x10<sup>-2</sup> | 0.03 | 0.011 - 0.08 |
| rs879043235 | 8137 | C>T | MT-CO2 | 4.278x10<sup>-12</sup> | 2.942x10<sup>-2</sup> | 1.085x10<sup>-2</sup> - 7.977x10<sup>-2</sup> | 0.03 | 0.011 - 0.08 |
| rs28358884 | 8414 | C>T | MT-ATP8 | 1.545x10<sup>-8</sup> | 1.029x10<sup>-1</sup> | 4.679x10<sup>-2</sup> - 2.263x10<sup>-1</sup> | 0.11 | 0.049 - 0.233 |
| rs201336180 | 8684 | C>T | MT-ATP6 | 2.706x10<sup>-12</sup> | 2.906x10<sup>-2</sup> | 1.078x10<sup>-2</sup> - 7.835x10<sup>-2</sup> | 0.03 | 0.011 - 0.079 |
| rs2298007 | 8794 | C>T | MT-ATP6 | 4.062x10<sup>-16</sup> | 2.247x10<sup>-4</sup> | 2.969x10<sup>-5</sup> - 1.7x10<sup>-3</sup> | 0.00 | 0 - 0.002 |
| rs3134801 | 9950 | T>C | MT-CO3 | 1.876x10<sup>-8</sup> | 3.782x10<sup>-2</sup> | 1.208x10<sup>-2</sup> - 1.184x10<sup>-1</sup> | 0.04 | 0.013 - 0.125 |
| rs878969753 | 10142 | C>T | MT-ND3 | 1.126x10<sup>-11</sup> | 3.074x10<sup>-2</sup> | 1.125x10<sup>-2</sup> - 8.401x10<sup>-2</sup> | 0.03 | 0.011 - 0.084 |
| rs41467651 | 10310 | G>A | MT-ND3 | 5.501x10<sup>-8</sup> | 6.296x10<sup>-3</sup> | 1.012x10<sup>-3</sup> - 3.916x10<sup>-2</sup> | 0.01 | 0.001 - 0.041 |
| rs28358278 | 10400 | C>T | MT-ND3 | 3.185x10<sup>-53</sup> | 2.007x10<sup>-2</sup> | 1.219x10<sup>-2</sup> - 3.306x10<sup>-2</sup> | 0.03 | 0.016 - 0.043 |
| rs28358283 | 10819 | A>G | MT-ND4 | 5.658x10<sup>-11</sup> | 1.809x10<sup>-1</sup> | 1.085x10<sup>-1</sup> - 3.017x10<sup>-1</sup> | 0.19 | 0.113 - 0.312 |
| rs2853497 | 12007 | G>A | MT-ND4 | 1.594x10<sup>-13</sup> | 1.277x10<sup>-1</sup> | 7.394x10<sup>-2</sup> - 2.206x10<sup>-1</sup> | 0.14 | 0.082 - 0.241 |
| rs878955011 | 12618 | G>A | MT-ND5 | 4.832x10<sup>-10</sup> | 1.08x10<sup>-1</sup> | 5.359x10<sup>-2</sup> - 2.177x10<sup>-1</sup> | 0.11 | 0.054 - 0.221 |
| rs386420001 | 12882 | C>T | MT-ND5 | 7.383x10<sup>-8</sup> | 4.087x10<sup>-3</sup> | 5.513x10<sup>-4</sup> - 3.029x10<sup>-2</sup> | 0.00 | 0.001 - 0.031 |
| rs2853501 | 13105 | A>G | MT-ND5 | 1.753x10<sup>-8</sup> | 1.796x10<sup>-1</sup> | 9.883x10<sup>-2</sup> - 3.263x10<sup>-1</sup> | 0.20 | 0.113 - 0.359 |
| rs879066842 | 13500 | T>C | MT-ND5 | 6.433x10<sup>-10</sup> | 9.579x10<sup>-2</sup> | 4.552x10<sup>-2</sup> - 2.016x10<sup>-1</sup> | 0.10 | 0.046 - 0.204 |
| rs28359184 | 13928 | G>C | MT-ND5 | 3.003x10<sup>-8</sup> | 6.307x10<sup>-2</sup> | 2.373x10<sup>-2</sup> - 1.676x10<sup>-1</sup> | 0.07 | 0.025 - 0.177 |
| rs28357672 | 14212 | T>C | MT-ND6 | 1.757x10<sup>-10</sup> | 1.39x10<sup>-1</sup> | 7.586x10<sup>-2</sup> - 2.549x10<sup>-1</sup> | 0.15 | 0.079 - 0.264 |
| rs386420019 | 14569 | G>A | MT-ND6 | 2.422x10<sup>-12</sup> | 7.852x10<sup>-2</sup> | 3.854x10<sup>-2</sup> - 1.6x10<sup>-1</sup> | 0.08 | 0.039 - 0.163 |
| rs28357678 | 14668 | C>T | MT-ND6 | 1.056x10<sup>-8</sup> | 1.52x10<sup>-1</sup> | 7.973x10<sup>-2</sup> - 2.898x10<sup>-1</sup> | 0.16 | 0.083 - 0.299 |
| rs28357680 | 14783 | T>C | MT-CYB | 1.036x10<sup>-53</sup> | 2.274x10<sup>-2</sup> | 1.406x10<sup>-2</sup> - 3.677x10<sup>-1</sup> | 0.03 | 0.018 - 0.433 |
| rs193302985 | 15043 | G>A | MT-CYB | 9.625x10<sup>-59</sup> | 4.309x10<sup>-2</sup> | 2.943x10<sup>-2</sup> - 6.309x10<sup>-2</sup> | 0.06 | 0.039 - 0.082 |
| rs193302991 | 15301 | G>A | MT-CYB | 2.787x10<sup>-84</sup> | 2.61x10<sup>-2</sup> | 1.808x10<sup>-2</sup> - 3.769x10<sup>-2</sup> | 0.05 | 0.032 - 0.065 |
| rs2853817 | 16172 | T>C | MT-HV1 | 2.132x10<sup>-8</sup> | 3.969x10<sup>-1</sup> | 2.873x10<sup>-1</sup> - 5.485x10<sup>-1</sup> | 0.42 | 0.306 - 0.571 |
| rs35134837 | 16217 | T>C | MT-HV1 | 8.59x10<sup>-10</sup> | 3.452x10<sup>-2</sup> | 1.177x10<sup>-2</sup> - 1.012x10<sup>-1</sup> | 0.04 | 0.012 - 0.107 |
| rs138126107 | 16261 | C>T | MT-HV1 | 2.04x10<sup>-9</sup> | 3.492x10<sup>-1</sup> | 2.476x10<sup>-1</sup> - 4.926x10<sup>-1</sup> | 0.36 | 0.256 - 0.503 |
| rs879067317 | 16318 | A>T | MT-HV1 | 1.887x10<sup>-10</sup> | 3.008x10<sup>-2</sup> | 1.024x10<sup>-2</sup> - 8.841x10<sup>-2</sup> | 0.03 | 0.01 - 0.089 |

### 3.4 Annotation of ALS SNVs

Annotation of ALS SNVs reveals that the control region (D-loop), protein coding genes and ribosomal RNA variants are associated with ALS (Table 1 and Table 2). The SNVs that increase the odds of ALS (Table 1) are in the hypervariable regions 1, 2 and 3, small subunit ribosomal ribonucleic acid (SSU rRNA) of the mitochondrial ribosome, NADs 5 and 6, Cytochrome c oxidases 1 and 3 and Cytochrome B. One nonsynonymous SNV with *p-*value <10^−7^ and OR >1 is found in *ND6* at 14318 (Asp>Ser). Seven nonsynonymous SNVs with OR <1, are observed in *ND2* (4824 Thr>Ala and 5178 Leu>Met), *ATP8* (8414 Leu>Phe), *ATP6* (8684 Thr>Ile and 8794 His>Tyr) and *ND5* (13105 Ile>Val and 13928 Ser>Thr) (Table 2). All other ALS SNVs are either synonymous or located in noncoding regions.

### 3.5 Mitochondrial haplogroups and ALS SNVs

Distribution of haplogroups in cases and controls shows three main haplogroups, namely H, U and L (Figure 2C). Haplogroup H is associated with Western European lines of descent, haplogroup U is associated with European Mediterranean populations and haplogroup L is associated with African and Middle Eastern (L3) populations. Haplogroup H contains more cases than controls (1,016 *vs* 259), haplogroup U has approximately the same frequency of cases and controls (178 *vs* 187) and haplogroup L contains less cases than controls (95 *vs* 687) (Figure 2C).

Because these three haplogroups are the most frequent in the dataset we performed intra-haplogroup GWAS and the results are detailed in Table 3. In haplogroup H, all SNVs identified were also identified in the general GWAS (Tables 1 and 2). In haplogroup L no non-synonymous variants were observed, and only one variant is also present in the main GWAS (rs28358568) (Table 3). In haplogroup U three unique ALS-SNVs were identified by intra-haplogroup GWAS being 2 synonymous in genes *HV2* and *ND5* and one missense variant in *ND2* (5186 Trp>Cys) (Table 3). The small differences observed in intra-haplogroup versus inter-haplogroup analysis are due to fact that haplogroup H, that show the same SNPs in both analyses, have significantly more cases than controls whereas haplogroups L and U have more controls than cases (Figure 2C). The ethnicity of all samples and the ALS patients was verified using the admixture analysis (Figure S4) which confirms the bias towards European ancestry suggested by mitochondrial haplogroups.

**TABLE 3.** Intra haplogroup GWAS of SNVs associated with ALS. Only variants with *p*-value < 10^−5^ are shown. The *p*-values were calculated by the *X^2^* test. *MT* = mitochondrial, *HV2* = hyper variable region 2, *HV3* = hyper variable region 3, *ND* = NADH ubiquinone oxidoreductases, *MT-TT* = mitochondrially encoded tRNA threonine, *RNR1* = 12S ribosomal RNA and *RNR2* = 16S ribosomal RNA. Variants were annotated using the MITOMAP (https://www.mitomap.org/MITOMAP) numbers confirmed in dbSNP build 152 (www.ncbi.nlm.nih.gov/snp/). C.I. = 95% confidence interval of the Odds Ratio. Hap. = Haplogroup. (*) = the odds ratios and C.I. of these variants were calculated with Haldane-Anscombe correction. (**) ALS-associated variants also detected in the analysis of 4,512 samples dataset (from Table 1).

| Hap. | dbSNP ID | Position | Change | Gene | P-value | Odds Ratio | C.I. |
| --- | --- | --- | --- | --- | --- | --- | --- |
| L | - | 76 | C>T | *MT-HV2* | $1.175 \times 10^{-6}$ | 30.15 | 3.334 – 272.7 |
| L | rs879040416 | 198 | C>T | *MT-HV2* | $4.605 \times 10^{-19}$ | 99.18 | 12.73 – 772.5 |
| L | - | *517 | del A | *MT-HV3* | $3.06 \times 10^{-6}$ | 52.02 | 20.33 – 133.12 |
| L | rs28358568 | **710 | T>C | *MT-RNR1* | $2.518 \times 10^{-22}$ | 118.6 | 15.39 – 913.5 |
| L | rs28619217 | 2755 | A>G | *MT-RNR2* | $4.766 \times 10^{-7}$ | 15.37 | 3.778 – 62.54 |
| L | rs41423746 | *3348 | A>G | *MT-ND1* | $3.06 \times 10^{-6}$ | 52.02 | 20.33 – 133.12 |
| L | rs1556423778 | 10196 | C>T | *MT-ND3* | $3.175 \times 10^{-6}$ | 30.15 | 3.334 – 272.7 |
| L | rs1569484396 | *10793 | C>T | *MT-ND4* | $3.06 \times 10^{-6}$ | 52.02 | 20.33 – 133.12 |
| L | rs1603224091 | 13209 | C>T | *MT-ND5* | $1.417 \times 10^{-6}$ | 19.03 | 3.638 – 99.52 |
| L | - | 13608 | T>C | *MT-ND5* | $3.175 \times 10^{-6}$ | 30.15 | 3.334 – 272.7 |
| L | - | *15940 | del T | *MT-TT* | $8.239 \times 10^{-36}$ | 396.81 | 162.97 – 966.17 |
| U | rs368463610 | *234 | A>G | *MT-HV2* | $7.194 \times 10^{-6}$ | $2.229 \times 10^{-1}$ | $9.402 \times 10^{-3}$ – $5.558 \times 10^{-2}$ |
| U | rs878939965 | *5186 | A>T | *MT-ND2* | $6.1112 \times 10^{-6}$ | $1.96 \times 10^{-2}$ | $8.07 \times 10^{-3}$ – $4.77 \times 10^{-1}$ |
| U | rs1556424235 | 13194 | G>A | *MT-ND5* | $6.112 \times 10^{-6}$ | $4.028 \times 10^{-2}$ | $5.38 \times 10^{-3}$ – $3.017 \times 10^{-1}$ |

The frequency of haplogroups H, U and L in the most severe, bulbar onset, *versus* limb, spinal and axial onsets was calculated. For this analysis we used 1,460 ALS cases for which data on the site of ALS onset was available. In ALS cases, 50.68% belonged to haplogroup H, 9.04% to haplogroup U and 4.73% to haplogroup L. In patients with Bulbar onset, 46.15% belonged to haplogroup H, 9.72% to haplogroup U and 4.86% to haplogroup L (Table 4). This suggests that the proportion of Bulbar onset *versus* other sites of onset is approximately the same in these haplogroups (Table 4).

**Table 4.**
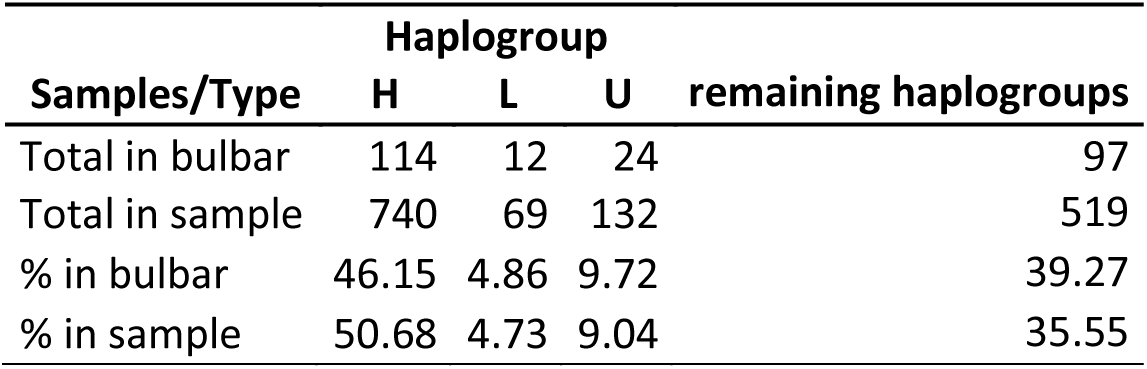
Frequency of bulbar ALS cases by haplogroup. The % in the sample corresponds to the total of cases per haplogroups the total of the sample. A total of 1.460 samples with clinical data were evaluated, 247 of which had a bulbar onset.

### 3.6 Alternative control datasets

To test if variations in the control dataset might affect the results, we produced two alternative datasets: (1) to verify haplogroup bias we generated one dataset with balanced haplogroup distribution by removal of 860 ALS samples to produce the dataset of 1,106 ALS samples, balanced for haplogroups, respective to the 2,546 control samples (Figures S5, S6 and S7) and (2) another dataset with a larger control mitochondrial representation encompassing the 17,736 unaffected mitogenomes analyzed by Wei and collaborators ^13^ (Figures S8, S9 and S10). Results with different control datasets reveal that the SNVs identified in association with ALS are constant especially when genes that contain SNPs that reduce the risk of ALS (Odds Ratios < 1) are compared (Tables S6 to S9) ^13^.

### 3.7 Heteroplasmy

Analysis of global heteroplasmy carried out on 1,978 mitogenomes (1,965 cases and 13 controls) shows that 26.7% of the sites found are heteroplasmic, and 73.3% are homoplasmic (Figure 3) ^29^. The distribution of mtDNA variants reveals specific profiles of allelic exchange frequencies per position (Figure 3), supported by high sequencing coverage (≈ 8,000x) (Figure 5). Homoplasmic and heteroplasmic sites are indicated along the mtDNA (Figure 3C), and coverage (Figure 3). The summary of heteroplasmy analysis is in Table S10.

**FIGURE 3.**
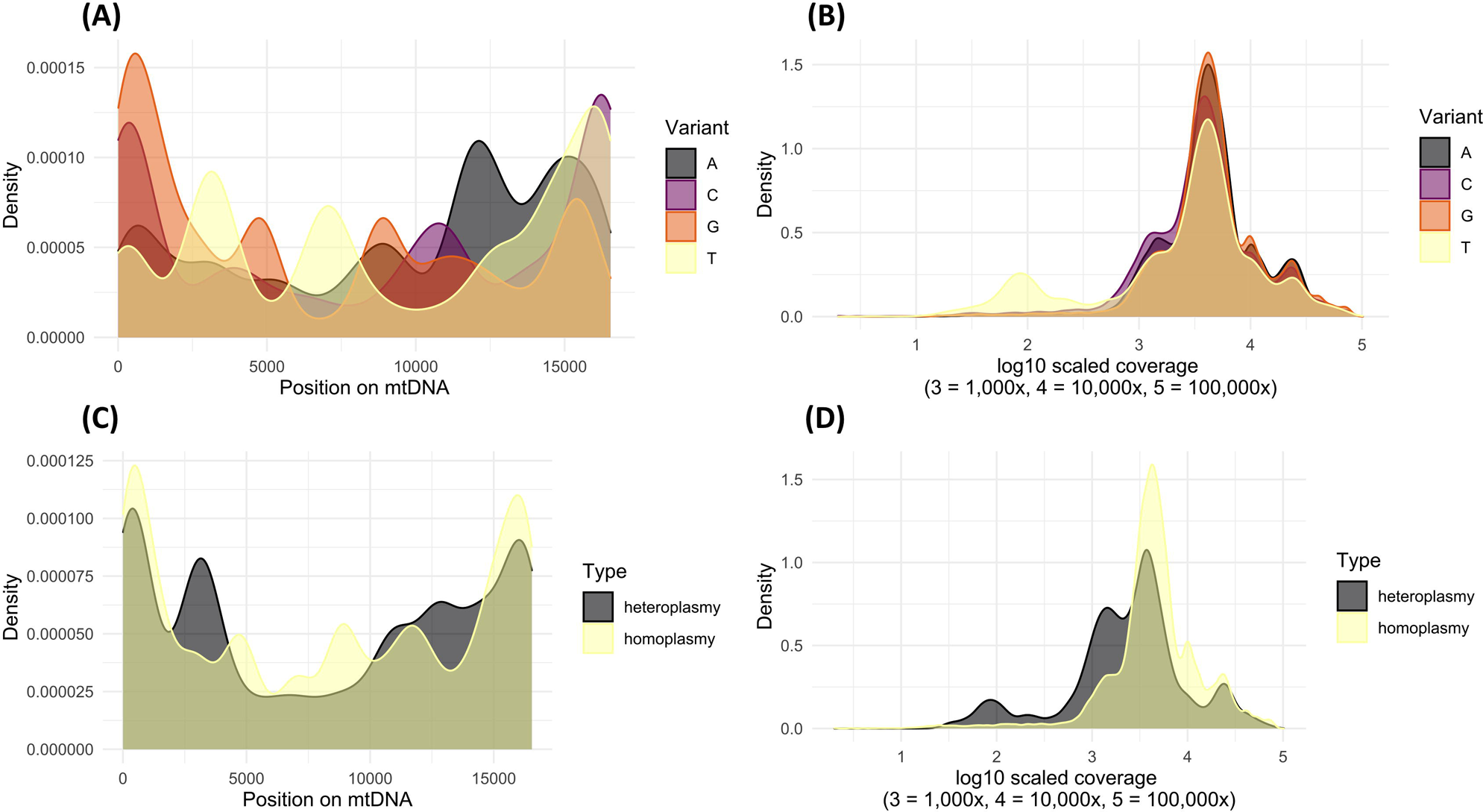
Heteroplasmy analysis (1,978 samples). Density plots show the distribution of variants by position (A), distribution of sequencing coverage by variant (B), distribution of variant type by position (C) and distribution of sequencing coverage by variant type (D).

### 3.8 Analysis of non-synonymous SNVs

Analysis of non-synonymous SNVs by PolyPhen ^30^ indicates that 14318 (aAt/aGt) is deleterious (FatHmm), 4824 (Acc/Gcc) 5178 (Cta/Ata) and 8414 (Ctc/Ttc) are possibly damaging (PolyPhen2) and deleterious (CADD and MtoolBox). SNVs 8684 (aCc/aTc), 8794 (Cac/Tac), 13105 (Atc/Gtc) and 13928 (aGc/aCc) are predicted to have either benign or neutral effects (Table S11). However, all are listed as benign by ClinVar.

Among variants that are associated with increased risk for ALS (Table 1), only one (rs28358587) has an association described for another phenotype (Resistance to high altitude pulmonary edema, HAPE), while 13 reduced risk variants (Table 2) show associations with other diseases (Table S12).

## 4 DISCUSSION

In this present study we observed, via GWAS, that at least eight nonsynonymous mutations in mitochondrion encoded genes could affect mitochondrial function and are associated with ALS with *p*-values <10^−7^ These nonsynonymous mutations led to two amino acid changes (m.4824A>G Thr>Ala, m.5178C>A Leu>Met) in *ND2*, two (m.1305A>G Ile>Val, m.13928G>C Ser>Thr) in *ND5*, one amino acid change (m.14318T>C Asn>Ser) in *ND6* (*ND2*, *ND5* and *ND6* are members of respiratory complex I), two changes (m.8684C>T Thr>Ile, m.8794C>T His>Tyr) in *ATP6* (complex V), and one change (m.8414C>T Leu>Phe) in *ATP8* (also complex V) (Table 1 and Table 2).

The total number of SNVs associated with ALS in this case-control sample is 51 for *p*-values <10^−7^. This threshold is extremely low considering the size of mitogenome (16,569bp) of approximately 1.6×10^4^ and the order of SNVs tested is 10^3^ (1,946 SNVs) which would implicate that a strict Bonferroni correction would draw the threshold around *p*-values below 10^−4^. The effective number of mitochondrial SNVs associated with ALS would therefore be higher. However, the threshold used here, with logistic regression, is conservative to mitigate false positives. The SNVs selected are well above the expected positive association by sheer chance due to multiple tests (Figure 2A). Also, the threshold of 10^−7^ would be used in case mtDNA was analyzed together with the nuclear genome in the same GWAS. Therefore the 10^−7^ is used so that it is possible to infer that the 51 SNVs here selected would still be statistically associated with ALS in a whole genome analysis.

Sequences in the control set of the main association analysis (Figure 2B) are the low-coverage whole genome sequencing (lcWGS) from 1KGP phase 3 ^31^. While lcWGS data returns a low nuclear DNA coverage (usually around 1x to 2x), it still includes a high mtDNA coverage due to the much higher copy number of mtDNA, which is present at hundreds or thousands of copies per cell ^32^. The mean mt DNA coverage for each individual ranged from 37.7x to 3,535x ^33^. Therefore, while standard variant calling on the low-coverage nuclear DNA is challenging and prone to errors, reliable variant calling in the mitochondrial genome is feasible because the mtDNA read depth in the same sample is hundreds to thousands of times higher. In fact, lcWGS has been used to analyze mtDNA as part of the 1KGP ^33^, as well as phylogenetic analysis of mitogenomes ^34^. Also, a usual concern in using the 1KGP sequences as controls in association studies focuses on the problem of imputation ^35^, which was not an issue in the analysis presented here.

Different control groups were used to test haplogroup bias and wider representation. Analyses with both sets supported conclusions of the main analysis. The differences between haplogroups in cases and controls indicate that mutations that simply differentiate the haplogroups are not artificially inflating the ALS associated SNVs. In intra-haplogroup analysis the SNVs associated with ALS are the same as compared to general GWAS. Because H is the most abundant haplogroup in ALS cases, at least in the population here tested, the SNVs below the *p*-value threshold of 10^−7^ still hold. As the sample number decreases, the *p-*values increase and that is why in U and L fewer SNVs are positive for ALS association, most likely because the sample is smaller (Tables 1, 2 and 3). To evidence the impact of population structure in our analysis, the odds ratios obtained if the population was unstructured are depicted in Supplementary Table S1. For example, in the case of rs28358875 (mtDNA position 7196) the odds ratio without correction is OR = 0.2725 by either Chi-Square or Fisher’s exact test (Table S5), and with correction by logistic regression, OR = 136.5 (Table 1). The differences in results obtained with different control datasets are a general characteristic of GWAS, that might lead to false positives depending on the cutoff values considered. Corrections for population heterogeneity and structure mitigate these false positives ^36^.

It is noteworthy that 30% of SNVs that increase the risk of ALS are in the control region (D-loop variable regions 1 and 2) while 16% of protective SNVs are in this region. It is therefore possible that besides direct effects in respiratory complex subunits, ALS mitochondria might be affected by variants associated with mtDNA replication.

A previous study of ALS patients from mainland China reported that ALS patients have a higher mean number of variants in protein-coding genes *ND4L*, *ND5*, *ND6*, and *ATP8* and that haplogroups Y and M7c may modulate ALS clinical expression ^12^. However, this conclusion was based on the analysis of whole mitochondrial genomes of only thirty-eight ALS patients and 42 unaffected controls. Moreover, their haplogroup classification (585 ALS patients and 371 healthy controls), considered only mitochondrial D-loop sequences although important substitutions that characterize several haplogroups occur outside the D-loop. Finally, Ni et al. ^12^ did not perform PCA to correct for populational structure. In our present study we found ALS associated SNPs in genes *ND4*, *ND5*, *ND6*, and *ATP8* (Tables 1 and 2), however, due to the limitations of the Ni et al. study ^12^, we cannot verify if SNPs described in our work coincide with variants they observed. Regarding haplogroups, both Y and M are significantly underrepresented in our ALS patients (Figure 2C), which precludes the verification that haplogroups Y and M7c modulate ALS clinical expression.

Potential limitations of our study include: (1) the majority of the cohort consists of individuals of European ancestry, (2) diagnosis was made at multiple sites and so may be subjected to variability, (3) Samples from multiple tissue types were included in the study, (4) potential technical confounders such as the use of both PCR free and PCR library preparations in sequencing, and (5) a replication cohort was not included.

Sufficient statistical data has been presented to suggest that these variants could be included in panels for ALS testing along with nuclear genome variants. Physical association between gene products of the mitogenome and the nuclear genome are essential for the formation of respiratory complexes I, III, IV and V (mitonuclear compatibility) ^37^. Although nonsynonymous mutations associated with ALS were present in subunits of complex I, III and V, synonymous mutations associated with ALS were identified and exert effects at the mRNA level and not only at the level of translated products. These synonymous SNVs might affect mRNA stability, association with mitochondrial rRNAs and tRNAs and impose selective constraints on mitochondrial codon usage.

## AUTHOR CONTRIBUTIONS

**Marcelo R. S. Briones:** Conceptualization; investigation; methodology; formal analysis; data curation; writing – original draft. **João H. Campos:** Investigation; methodology; formal analysis; data curation; visualization; editing – original draft. **Renata C. Ferreira**: Conceptualization; supervision; writing – review and editing. **Lisa Schneper:** Investigation; methodology; validation; data curation; supervision; writing – original draft. **Ilda M. Santos:** Investigation; methodology; validation; data curation. **Fernando M. Antoneli:** Investigation; methodology; formal analysis; statistical analysis; writing – original draft. **James R. Broach:**

Conceptualization; investigation; methodology; formal analysis; data curation; writing – original draft.

## ACKNOWLEDGEMENTS

The authors thank Fabricio Landi for assistance with computational facilities. J.H.C. was supported by a PhD fellowship from CAPES, I.M.S. was supported by an MSc fellowship from CAPES. This work was supported by grants to M. R. S. B. (FAPESP 2014/25602-6, FAPESP 2013/07838-0 and CNPq 303912/2017-0) and NIH grants to J. R. B. (#). All NYGC ALS Consortium activities are supported by the ALS Association (ALSA, 19-SI-459) and the Tow Foundation.

## DATA AVAILABILITY STATEMENT

All data produced in the present study are available upon reasonable request to the corresponding authors.

## ETHICS STATEMENT

We confirm that we have read the Journal’s position on issues involved in ethical publication and affirm that this report is consistent with those guidelines.

