## Supplementary material for "Mitochondrial genome variants associated with Amyotrophic Lateral Sclerosis and their haplogroup distribution": Figures S1-S10.docx

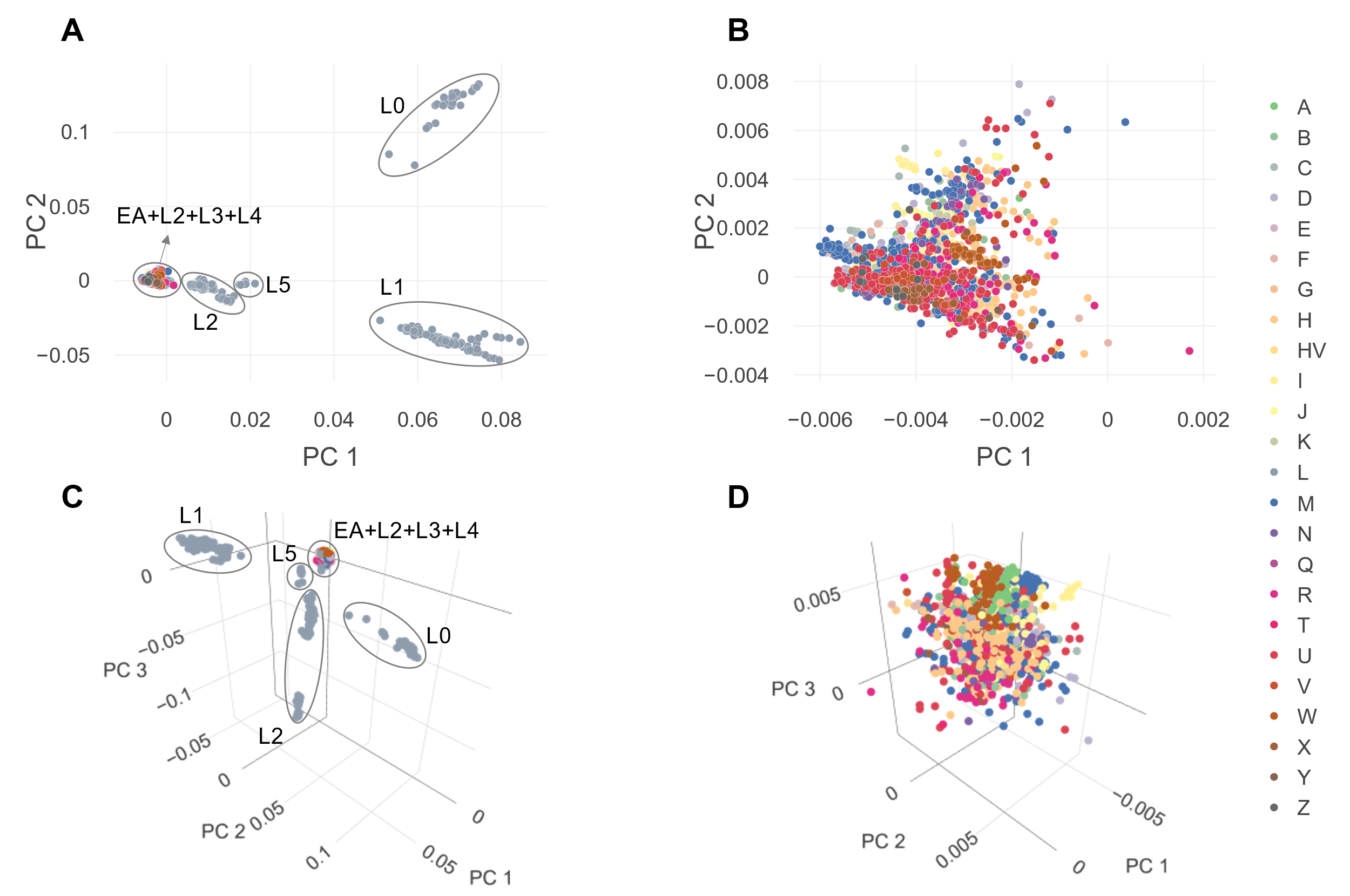

**Figure S1**. Principal Component Analysis (PCA) by haplogroup (4,512 samples). The color scale indicates the mitochondrial haplogroups. (A) the 2D PCA of 2,547 controls and 1,965 cases, (B) a closer look at the non-African cluster, (C) 3D PCA with a third component of all samples, and (D) the detailed 3D zoom in of the European-Asian haplogroups. African (L) and Eurasian (EA) haplogroups are highlighted (ovals) in A and B.

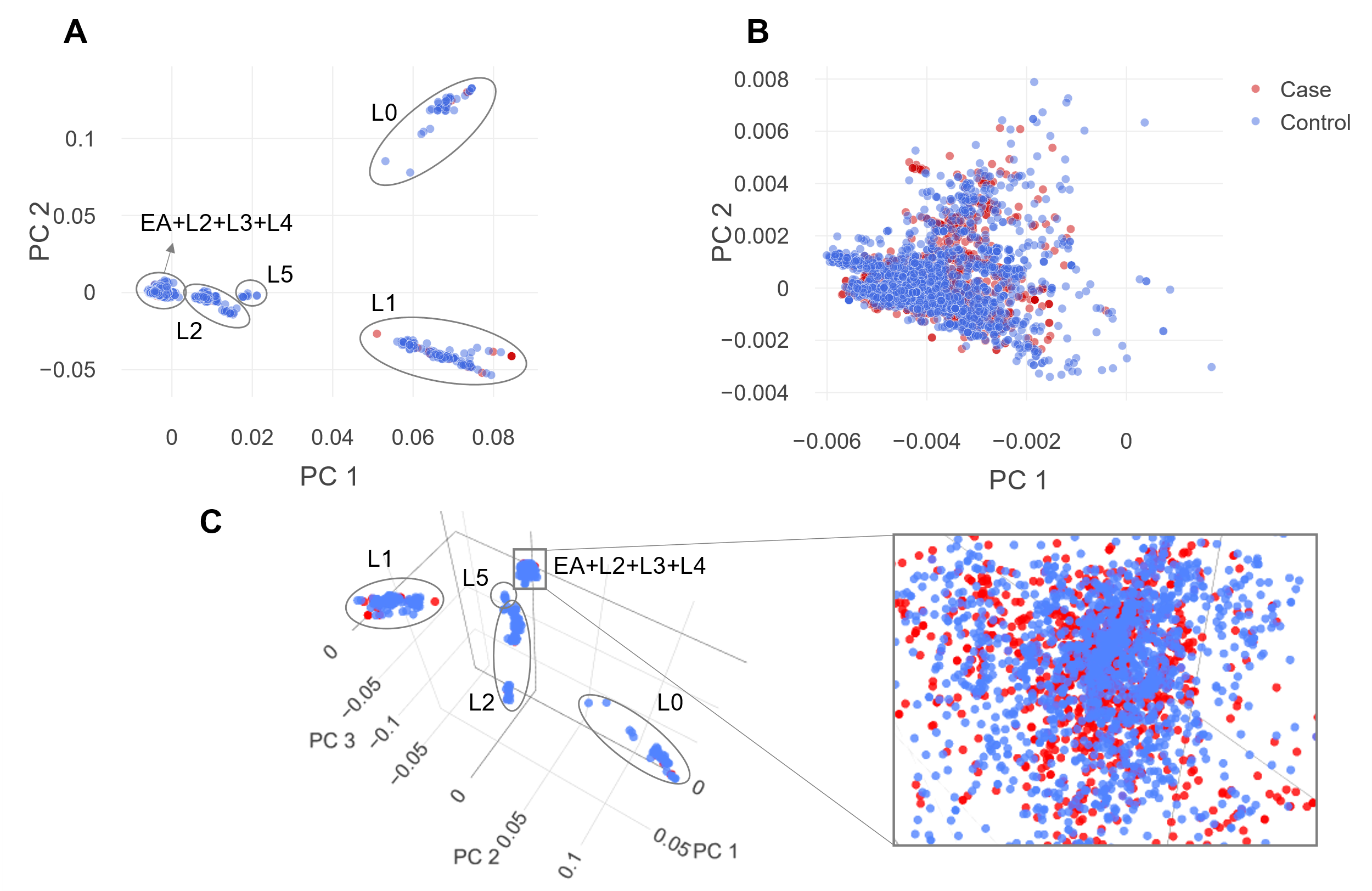

**Figure S2**. Principal Component Analysis (PCA) by group. 4,512 samples are represented in two groups: 2,547 controls (blue circles), and 1,965 cases (red circles) (A). The region that concentrates the largest number of samples is enlarged in B and details the overlapping of cases and controls. In C, the third component changes the perspective observed in A, and the highlighted box on the right further expands the sample distribution view. African (L) and Eurasian (EA) haplogroups are highlighted (ovals) in A and B.

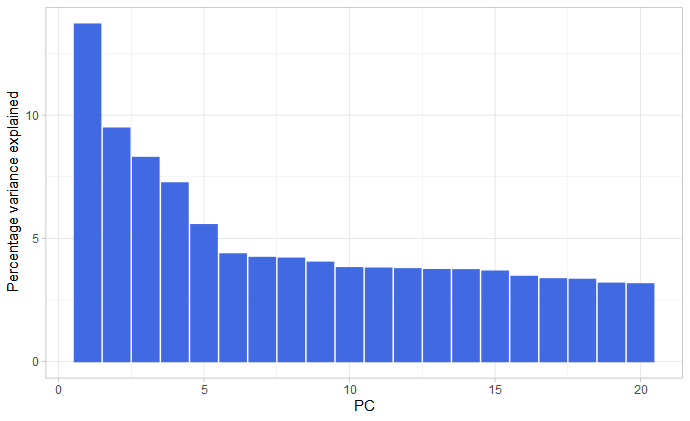

**Figure S3.** Scree plot showing the 20 components used in logistic regression. The components (horizontal axis) are depicted in decreasing order according to the percentage of variance explained (vertical axis).

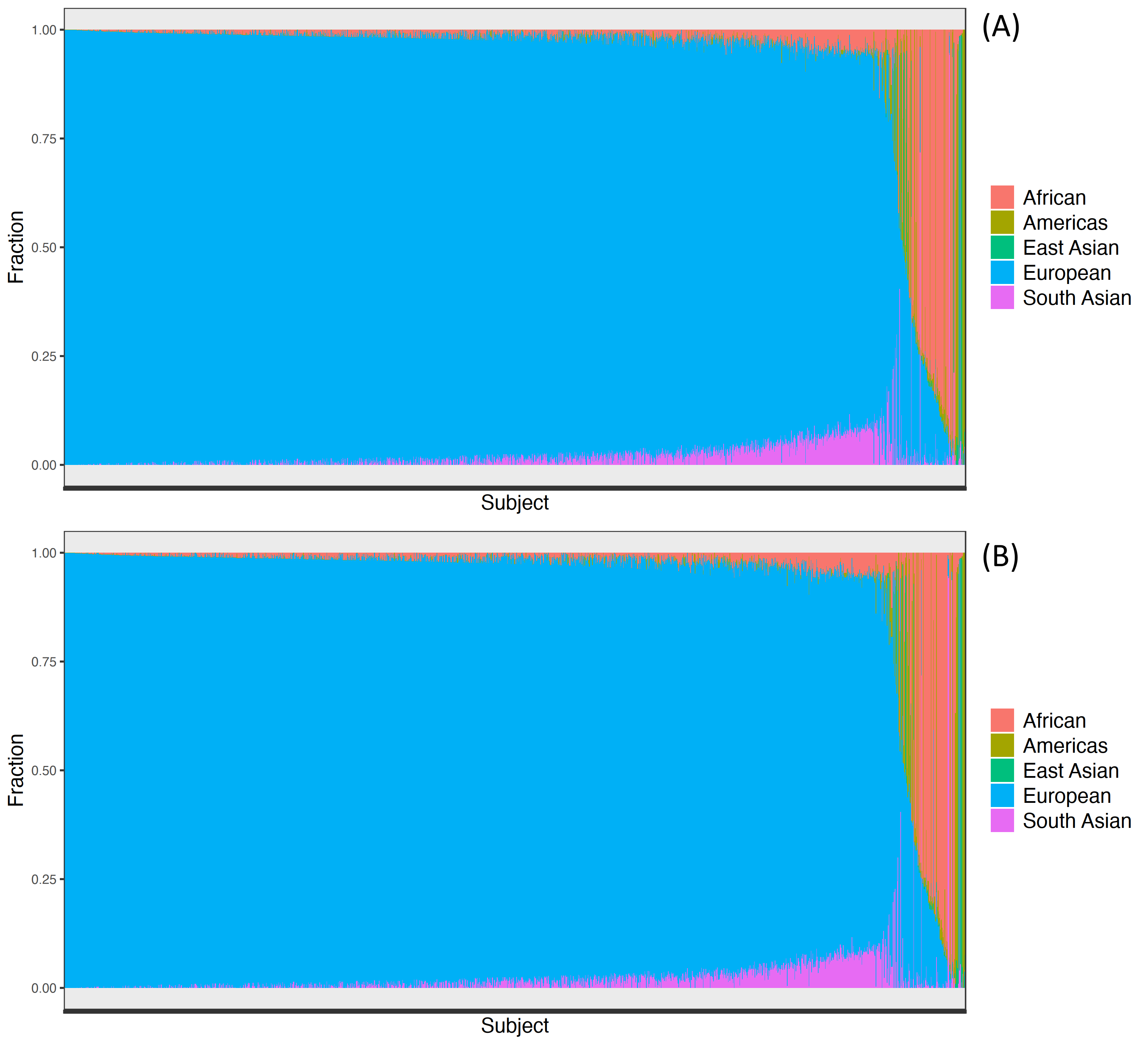

**Figure S4.** The ethnicity of samples used in the study was based on NYGC whole genome data. In (A) all samples and (B) only ALS samples. Panels represent the relative SNP contributions as determined by genome admixture analysis.

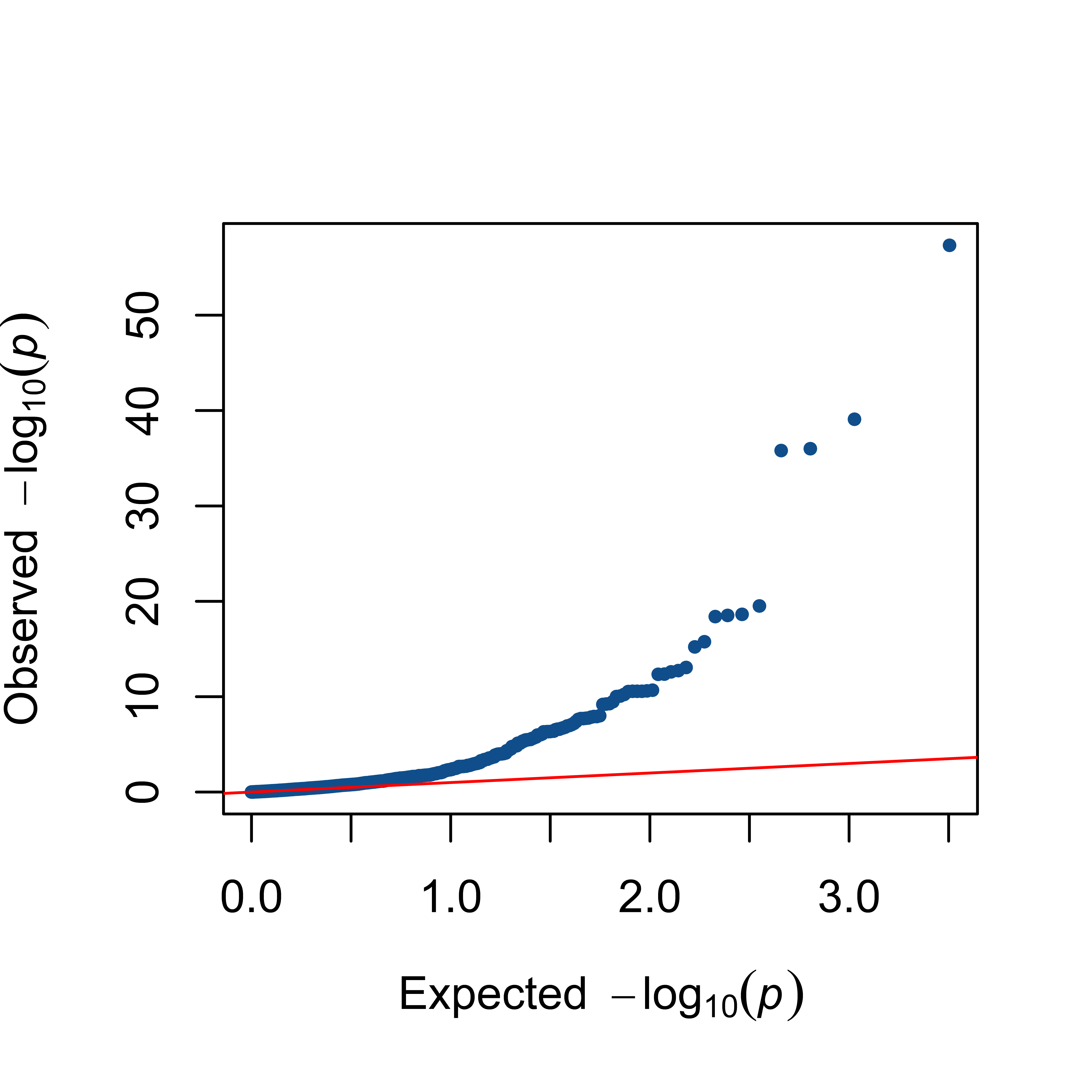

**Figure S5.** The QQ-Plot with logistic regression of the 1,934 SNVs selected after removal of SNVs with genotype failure rate >0.1 and Minimum Allele Frequency (MAF) <0.1%. The red line is the expected random association values. This corresponds to the analysis of 3,652 samples, being 1,106 cases and 2,546controls (for details see Material and Methods section).

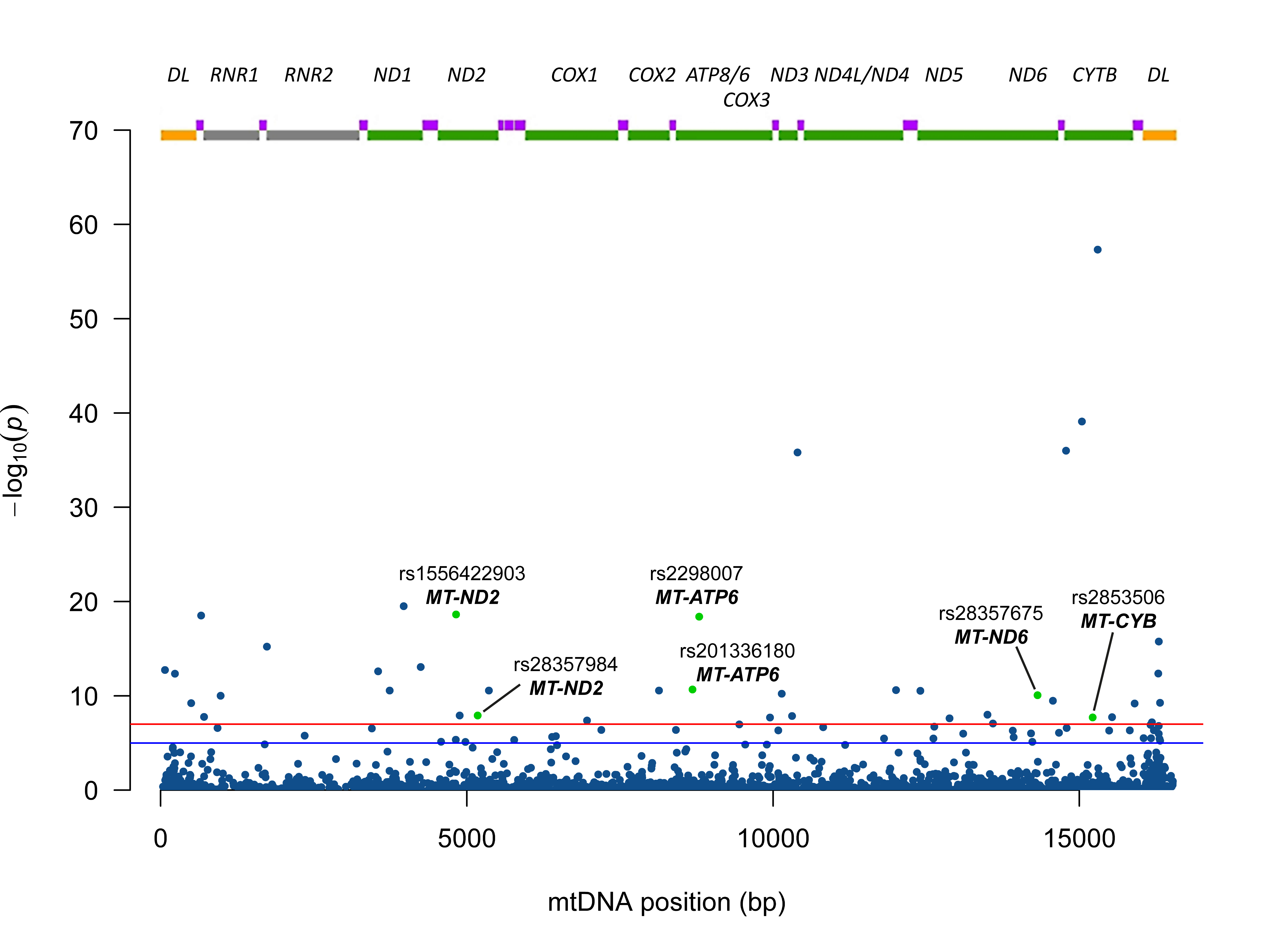

**Figure S6.** Manhattan plot of the 1,934 SNVs mapped to the mitochondrial genome. In the abscissa are the mitochondrial positions numbered according to the revised Cambridge Reference Sequence (rCRS) (30) and in the ordinate the -log10(p-values). The horizontal blue line indicates the 10^-5^ threshold and the red horizontal line the 10^-7^ threshold. The gene map is depicted at the top with yellow boxes representing the D-loop, green boxes the protein coding genes, gray boxes the rRNA genes and purple boxes the tRNA genes. This corresponds to the analysis of 3,652 samples, being 1,106 cases and 2,546 controls (for details see Material and Methods section). Nonsynonymous substitutions are depicted as green dots and annotated.

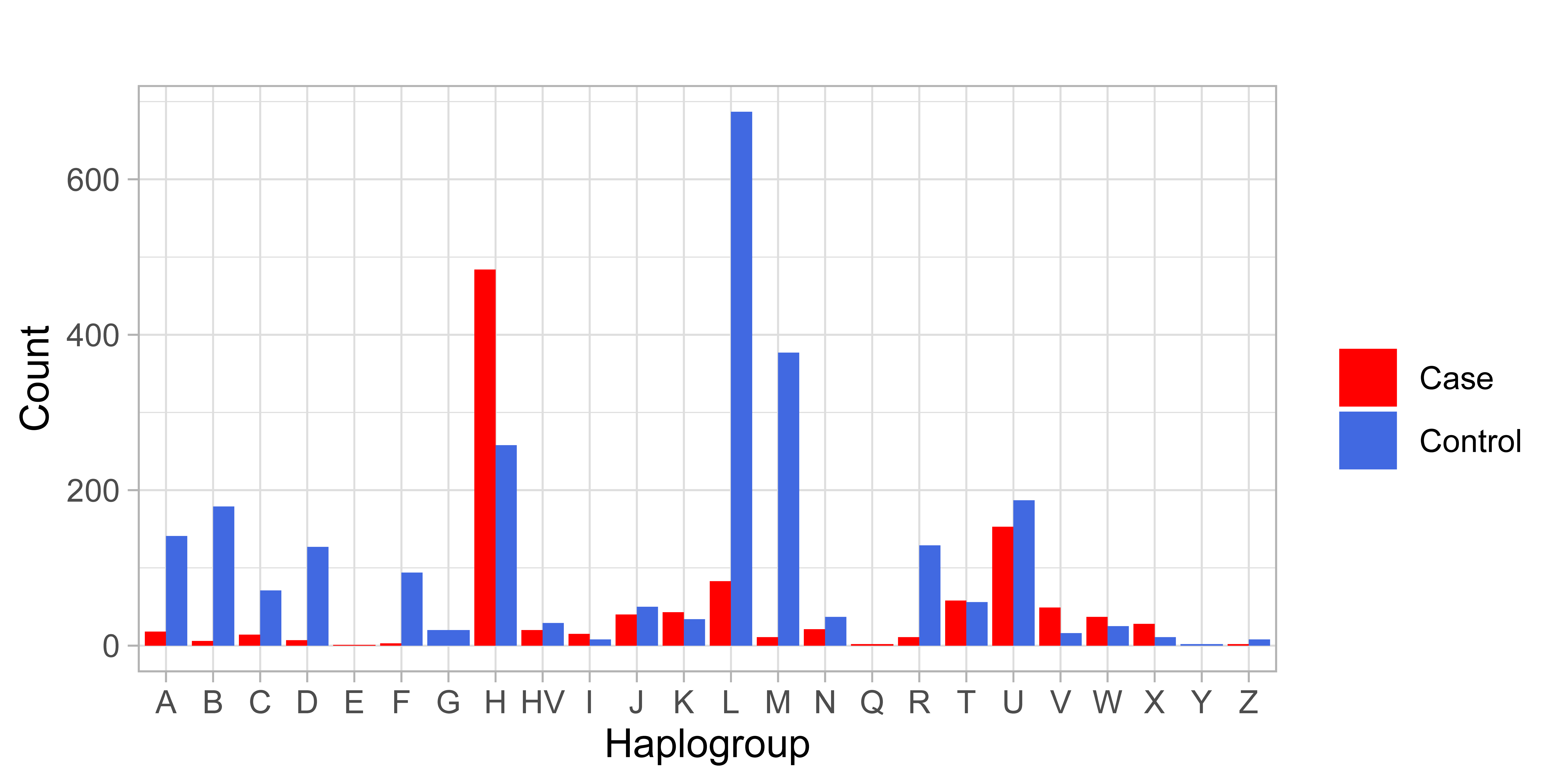

**Figure S7.** Plot of the mitochondrial haplogroup frequencies in 1,106 cases and 2,546 controls. The haplogroups are depicted in the abscissa and the number of samples in each category are depicted in the ordinate.

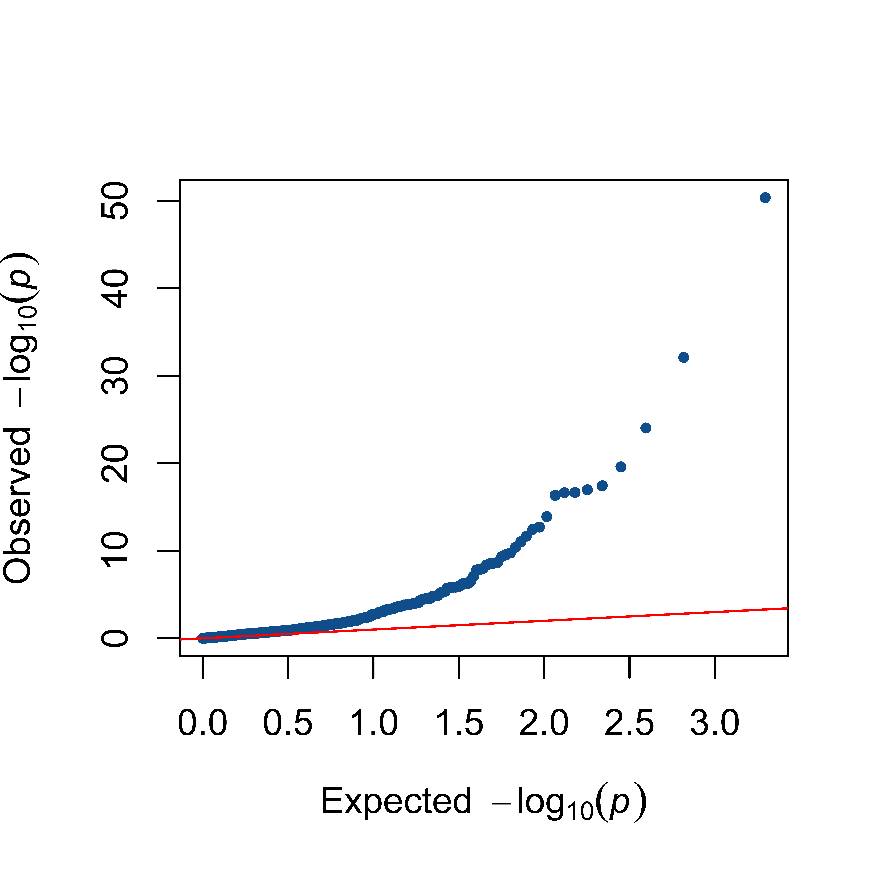

**Figure S8.** The QQ-Plot with logistic regression of the 1,011 SNVs selected after removal of SNVs with genotype failure rate >0.1 and Minimum Allele Frequency (MAF) <0.1%. The red line is the expected random association values. This corresponds to the analysis of 19,701 samples, being 1,965 cases and 17,736 controls (for details see Material and Methods section).

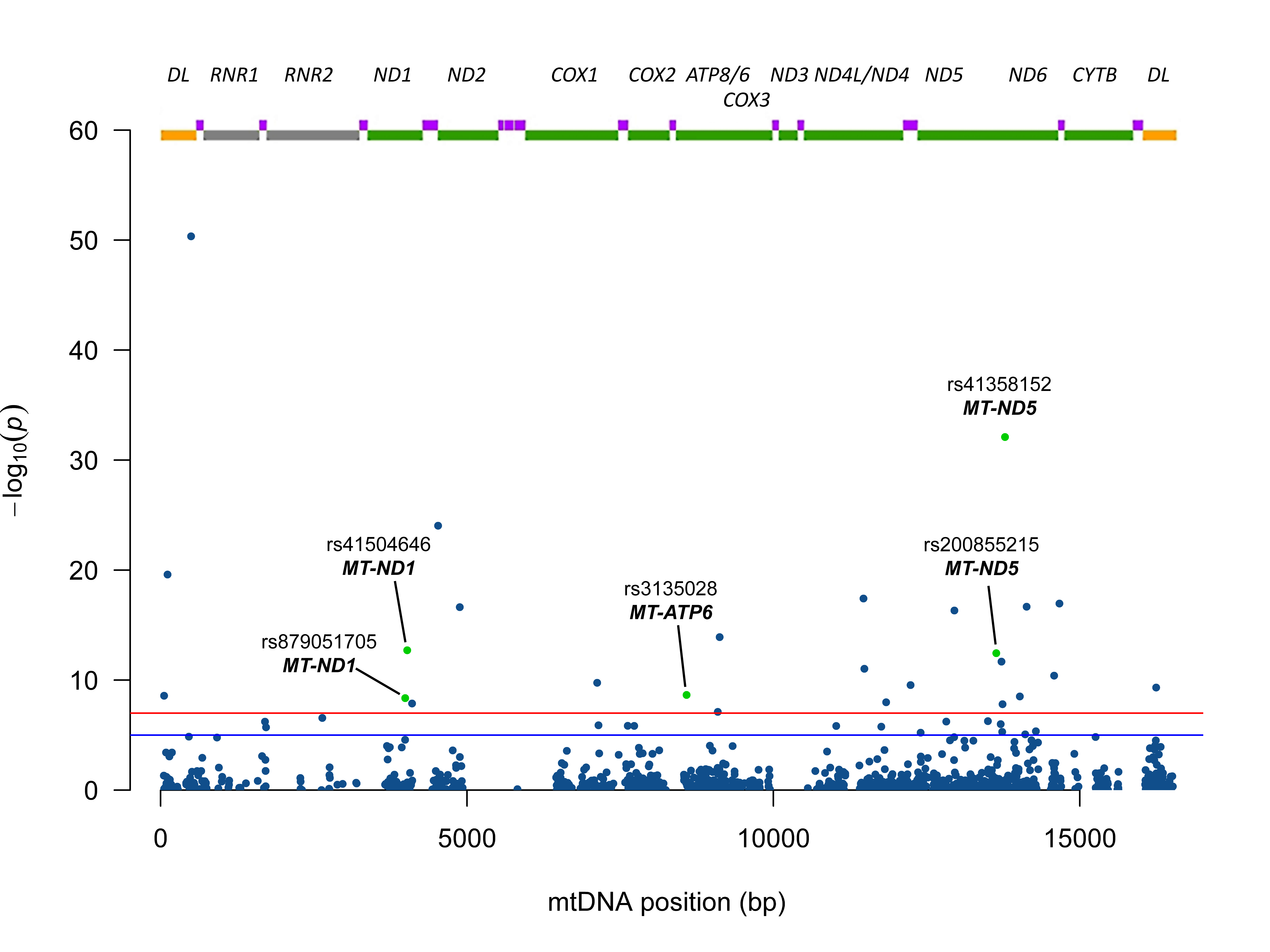

**Figure S9.** Manhattan plot of the 1,011 SNVs mapped to the mitochondrial genome. In the abscissa are the mitochondrial positions numbered according to the revised Cambridge Reference Sequence (rCRS) (30) and in the ordinate the -log10(p-values). The horizontal blue line indicates the 10^-5^ threshold and the red horizontal line the 10^-7^ threshold. The gene map is depicted at the top with yellow boxes representing the D-loop, green boxes the protein coding genes, gray boxes the rRNA genes and purple boxes the tRNA genes. This corresponds to the analysis of 19,701 samples, being 1,965 cases and 17,736 controls (for details see Material and Methods section). Nonsynonymous substitutions are depicted as green dots and annotated.

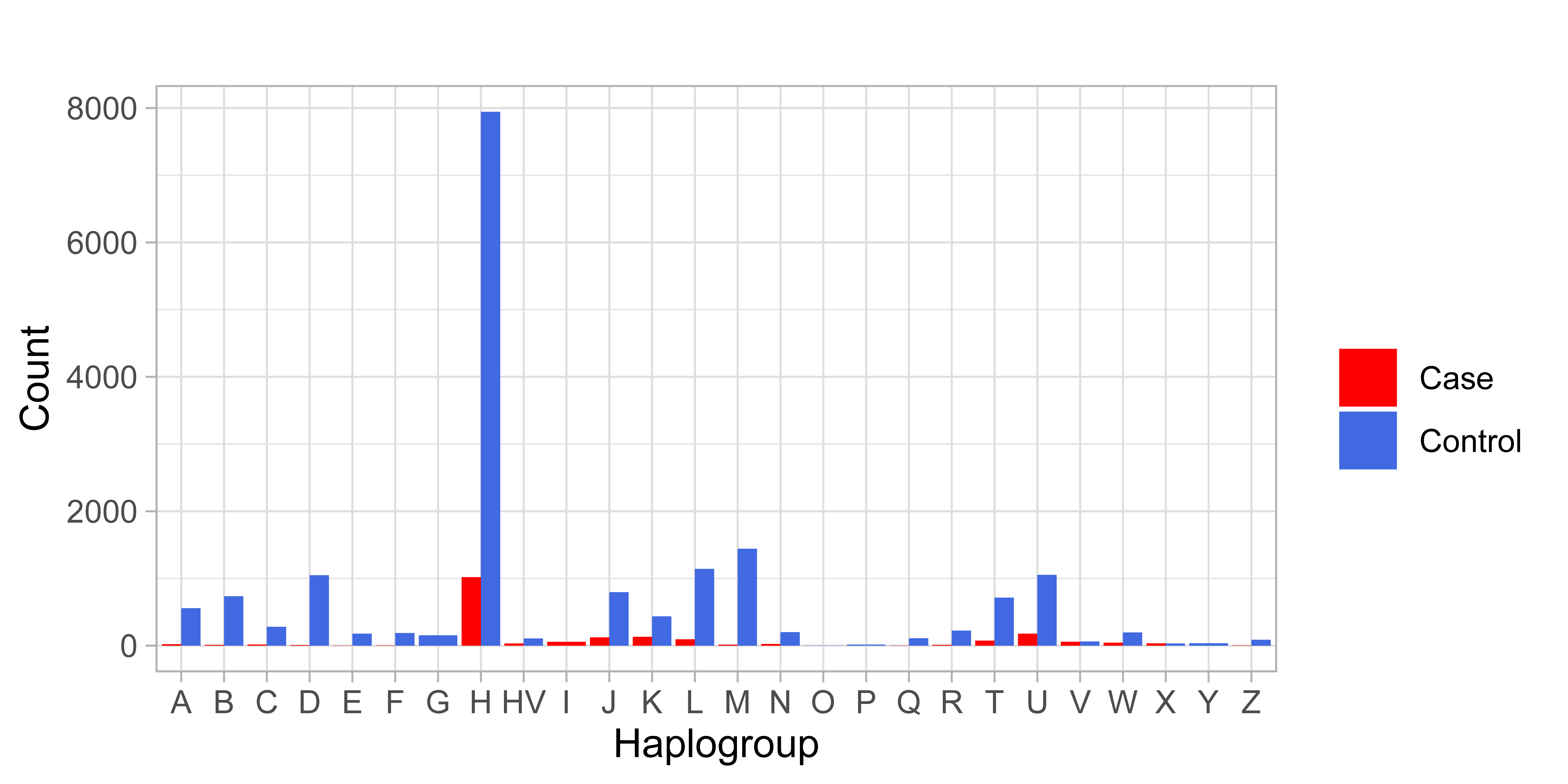

**Figure S10.** Plot of the mitochondrial haplogroup frequencies in 1,965 cases and 17,736 controls. The haplogroups are depicted in the abscissa and the number of samples in each category are depicted in the ordinate.
