## Supplementary material for "Mitochondrial genome variants associated with Amyotrophic Lateral Sclerosis and their haplogroup distribution": Table S6 to S9-Alternative controls.docx

**Supplementary Data**

**Table S6.** Mitochondrial genome variants that increase the risk of ALS in the haplogroups balanced dataset. Only variants with p-value <10^-7^. Logistic regression of 2,546 controls *vs* 1,106 ALS cases (3,652 samples). *MT* = mitochondrial, *HV* = hyper variable region*, MT-TT* = *tRNA threonine*, *CYB* = Cytochrome *b*, *ND* = NADH ubiquinone oxidoreductases*, RNR1*= 12S ribosomal RNA. Nucleotide changes as shown in PLINK program. Variants annotated using NCBI Variation Viewer. (<https://www.ncbi.nlm.nih.gov/variation/view/>). C.I. = 95% confidence interval of the Odds Ratio.

| **dbSNP ID** | **Position** | **Change** | **Gene** | ***P-*Value** | **Odds Ratio** | **95% C.I.** |
| --- | --- | --- | --- | --- | --- | --- |
| rs879158303 | 72 | T>C | *MT-HV2* | 1.842x10^-13^ | 6.597 | 3.992 - 10.9 |
| rs28358568 | 710 | T>C | *MT-RNR1* | 1.725x10^-8^ | 290.6 | 40.45 - 2088 |
| rs28358587 | 3552 | T>A | *MT-ND1* | 2.55x10^-13^ | 1217 | 181.5 - 8162 |
| rs1556424102 | 12405 | C>T | *MT-ND5* | 2.916x10^-11^ | 9.235x10^4^ | 3178 - 2.684x10^6^ |
| rs28357675 | 14318 | T>C | *MT-ND6* | 8.614x10^-11^ | 504.1 | 76.96 - 3301 |
| rs2853506 | 15218 | A>G | *MT-CYB* | 1.976x10^-8^ | 3.406 | 2.22 - 5.224 |
| rs35788393 | 15904 | C>T | *MT-TT* | 6.584x10^-10^ | 5.818 | 3.327 - 10.17 |
| rs879166752 | 16186 | C>T | *MT-HV1* | 6.417x10^-8^ | 5.905 | 3.102 - 11.24 |
| rs148377232 | 16298 | T>C | *MT-HV1* | 1.746x10^-16^ | 6.767 | 4.294 - 10.67 |

**Table S7.** Mitochondrial genome variants that reduce the risk of ALS in the haplogroup balanced dataset. Only variants with p-value <10^-7^. Logistic regression of 2,546 controls *vs* 1,106 ALS cases (3,652 samples). *MT* = mitochondrial, *ATP6* = ATP synthase subunit 6, *HV* = hyper variable region*, MT-TT* = *tRNA threonine*, *CO* = Cytochrome *c* oxidases, *CYB* = Cytochrome *b*, *ND* = NADH ubiquinone oxidoreductases*, RNR1*= 12S ribosomal RNA and *RNR2* = 16S ribosomal RNA. Nucleotide changes as shown in PLINK program. Variants annotated using NCBI Variation Viewer. (<https://www.ncbi.nlm.nih.gov/variation/view/>). C.I. = 95% confidence interval of the Odds Ratio.

| **dbSNP ID** | **Position** | **Change** | **Gene** | ***P-*Value** | **Odds Ratio** | **95% C.I.** |
| --- | --- | --- | --- | --- | --- | --- |
| rs3937037 | 235 | A>G | *MT-HV2* | 4.549x10^-13^ | 6.959x10^-4^ | 9.717x10^-5^ - 4.983x10^-3^ |
| rs3901846 | 499 | G>A | *MT-HV3* | 6.023x10^-10^ | 1.052x10^-2^ | 2.488x10^-3^ - 4.45x10^-2^ |
| rs56489998 | 663 | A>G | *MT-RNR1* | 3.039x10^-19^ | 7.152x10^-5^ | 8.879x10^-6^ - 5.761x10^-4^ |
| rs397515731 | 980 | T>C | *MT-RNR1* | 9.855x10^-11^ | 2.631x10^-2^ | 8.74x10^-3^ - 7.921x10^-2^ |
| rs193303006 | 1736 | A>G | *MT-RNR2* | 6.155x10^-16^ | 2.405x10^-4^ | 3.191x10^-5^ - 1.813x10^-3^ |
| rs878907222 | 3741 | C>T | *MT-ND1* | 2.757x10^-11^ | 1.915x10^-2^ | 5.977x10^-3^ - 6.134x10^-2^ |
| rs9629042 | 3970 | C>T | *MT-ND1* | 3.125x10^-20^ | 1.134x10^-7^ | 3.779x10^-9^ - 3.404x10^-6^ |
| rs9326618 | 4248 | T>C | *MT-ND1* | 8.756x10^-14^ | 5.4x10^-4^ | 7.477x10^-5^ - 3.9x10^-3^ |
| rs1556422903 | 4824 | A>G | *MT-ND2* | 2.365x10^-19^ | 6.936x10^-5^ | 8.607x10^-6^ - 5.589x10^-4^ |
| rs200763872 | 4883 | C>T | *MT-ND2* | 1.213x10^-8^ | 8.711x10^-2^ | 3.762x10^-2^ - 2.017x10^-1^ |
| rs28357984 | 5178 | C>A | *MT-ND2* | 1.213x10^-8^ | 8.711x10^-2^ | 3.762x10^-2^ - 2.017x10^-1^ |
| rs879217723 | 5360 | C>T | *MT-ND2* | 2.757x10^-11^ | 1.915x10^-2^ | 5.977x10^-3^ - 6.134x10^-2^ |
| rs1970771 | 6962 | G>A | *MT-CO1* | 4.156x10^-8^ | 2.479x10^-3^ | 2.905x10^-4^ - 2.116x10^-2^ |
| rs879043235 | 8137 | C>T | *MT-CO2* | 2.757x10^-11^ | 1.915x10^-2^ | 5.977x10^-3^ - 6.134x10^-2^ |
| rs201336180 | 8684 | C>T | *MT-ATP6* | 2.131x10^-11^ | 1.914x10^-2^ | 6.012x10^-3^ - 6.091x10^-2^ |
| rs2298007 | 8794 | C>T | *MT-ATP6* | 4.061x10^-19^ | 7.535x10^-5^ | 9.391x10^-6^ - 6.046x10^-4^ |
| rs3134801 | 9950 | T>C | *MT-CO3* | 2.013x10^-8^ | 9.917x10^-3^ | 1.979x10^-3^ - 4.969x10^-2^ |
| rs878969753 | 10142 | C>T | *MT-ND3* | 5.975x10^-11^ | 1.998x10^-2^ | 6.191x10^-3^ - 6.451x10^-2^ |
| rs41467651 | 10310 | G>A | *MT-ND3* | 1.386x10^-8^ | 1.03x10^-3^ | 9.576x10^-5^ - 1.108x10^-2^ |
| rs28358278 | 10400 | C>T | *MT-ND3* | 1.567x10^-36^ | 2.791x10^-2^ | 1.601x10^-2^ - 4.865x10^-2^ |
| rs2853497 | 12007 | G>A | *MT-ND4* | 2.49x10^-11^ | 7.285x10^-2^ | 3.375x10^-2^ - 1.572x10^-1^ |
| rs386420001 | 12882 | C>T | *MT-ND5* | 2.421x10^-8^ | 2.196x10^-3^ | 2.557x10^-4^ - 1.886x10^-2^ |
| rs879066842 | 13500 | T>C | *MT-ND5* | 1.003x10^-8^ | 7.29x10^-2^ | 2.977x10^-2^ - 1.785x10^-1^ |
| rs28359177 | 13590 | G>A | *MT-ND5* | 8.588x10^-8^ | 1.225x10^-2^ | 2.528x10^-3^ - 6.233x10^-2^ |
| rs386420019 | 14569 | G>A | *MT-ND6* | 3.365x10^-10^ | 5.628x10^-2^ | 2.293x10^-2^ - 1.381x10^-1^ |
| rs28357680 | 14783 | T>C | *MT-CYB* | 1.009x10^-36^ | 3.253x10^-2^ | 1.914x10^-2^ - 5.528x10^-2^ |
| rs193302985 | 15043 | G>A | *MT-CYB* | 8.209x10^-40^ | 5.911x10^-2^ | 3.885x10^-2^ - 8.995x10^-2^ |
| rs193302991 | 15301 | G>A | *MT-CYB* | 4.844x10^-58^ | 3.417x10^-2^ | 2.263x10^-2^ - 5.159x10^-2^ |
| rs28357371 | 15535 | C>T | *MT-CYB* | 1.838x10^-08^ | 8.74x10^-4^ | 7.519x10^-5^ - 1.016x10^-2^ |
| rs1556424849 | 16290 | C>T | *MT-HV1* | 4.319x10^-13^ | 6.579x10^-4^ | 9.065x10^-5^ - 4.774x10^-3^ |
| rs879067317 | 16318 | A>T | *MT-HV1* | 5.488x10^-10^ | 1.754x10^-2^ | 4.889x10^-3^ - 6.291x10^-2^ |

**Table S8.** Mitochondrial genome variants that increase the risk of ALS in the large Wei et al. (2017) control dataset. Only variants with p-value <10^-7^. Logistic regression of 17,736 controls *vs* 1,965 ALS cases (19,701 samples). *MT* = mitochondrial, *HV* = hyper variable region, *CO* = Cytochrome *c* oxidases and *ND* = NADH ubiquinone oxidoreductases. Nucleotide changes as shown in PLINK program. Variants annotated using NCBI Variation Viewer. (<https://www.ncbi.nlm.nih.gov/variation/view/>). C.I. = 95% confidence interval of the Odds Ratio.

| **dbSNP ID** | **Position** | **Change** | **Gene** | ***P*-value** | **Odds Ratio** | **95% C.I.** |
| --- | --- | --- | --- | --- | --- | --- |
| rs879102890 | 114 | C>T | *MT-HV2* | 2.552x10^-20^ | 7.147 | 4.708 -10.85 |
| rs28660704 | 497 | C>T | *MT-HV3* | 4.587x10^-51^ | 5.689 | 4.535 - 7.136 |
| rs879051705 | 3992 | C>T | *MT-ND1* | 4.332x10^-9^ | 4.638 | 2.779 - 7.74 |
| rs41504646 | 4024 | A>G | *MT-ND1* | 1.955x10^-13^ | 11.55 | 6.014 - 22.16 |
| rs1117205 | 4104 | A>G | *MT-ND1* | 1.343x10^-08^ | 88.55 | 18.85 - 415.9 |
| rs1556422869 | 4529 | A>T | *MT-ND2* | 9.297x10^-25^ | 62.24 | 28.3 - 136.9 |
| rs1603220784 | 7124 | A>G | *MT-CO1* | 1.762x10^-10^ | 71.6 | 19.28 - 265.9 |
| rs878968914 | 11470 | A>G | *MT-ND4* | 3.836x10^-18^ | 36.87 | 16.33 - 83.24 |
| rs28529320 | 11485 | T>C | *MT-ND4* | 9.299x10^-12^ | 4.301 | 2.828 - 6.542 |
| rs28550734 | 11840 | C>T | *MT-ND4* | 1.039x10^-8^ | 10.4 | 4.664 - 23.18 |
| rs878875112 | 12954 | T>C | *MT-ND5* | 4.76x10^-17^ | 33.53 | 14.76 - 76.16 |
| rs200855215 | 13637 | A>G | *MT-ND5* | 3.598x10^-13^ | 11.44 | 5.93 - 22.06 |
| rs386829190 | 13722 | A>G | *MT-ND5* | 2.096x10^-12^ | 6.244 | 3.747 - 10.41 |
| rs28630861 | 13740 | T>C | *MT-ND5* | 1.567x10^-8^ | 9.35 | 4.308 - 20.29 |
| rs41358152 | 13780 | A>G | *MT-ND5* | 8.06x10^-33^ | 43.09 | 23.22 - 79.94 |
| rs879100848 | 14133 | A>G | *MT-ND5* | 2.15x10^-17^ | 10.61 | 6.147 - 18.3 |
| rs41354845 | 14582 | A>G | *MT-ND6* | 3.946x10^-11^ | 9.212 | 4.767 - 17.8 |

**Table S9.** Mitochondrial genome variants that reduce the risk of ALS in the large Wei et al. (2017) control dataset. Only variants with p-value <10^-7^. Logistic regression of 17,736 controls *vs* 1,995 ALS cases (19,701 samples). *MT* = mitochondrial, *ATP6* = ATP synthase subunit 6, *HV* = hyper variable region, *ND* = NADH ubiquinone oxidoreductases and *TS2* = mitochondrially encoded TRNA-Ser (AGU/C) 2. Nucleotide changes as shown in PLINK program. Variants annotated using NCBI Variation Viewer. (<https://www.ncbi.nlm.nih.gov/variation/view/>). C.I. = 95% confidence interval of the Odds Ratio.

| **dbSNP ID** | **Position** | **Change** | **Gene** | ***P*-value** | **Odds Ratio** | **95% C.I.** |
| --- | --- | --- | --- | --- | --- | --- |
| - | 58 | T>C | *MT-HV2* | 2.657x10^-9^ | 1.495x10^-4^ | 8.221x10^-6^ - 2.719x10^-3^ |
| rs200763872 | 4883 | C>T | *MT-ND2* | 2.35x10^-17^ | 4.889x10^-2^ | 2.432x10^-2^ - 9.825x10^-2^ |
| rs3135028 | 8584 | G>A | *MT-ATP6* | 2.257x10^-9^ | 2.414x10^-1^ | 1.515x10^-1^ - 3.847x10^-1^ |
| rs28358270 | 9123 | G>A | *MT-ATP6* | 1.25x10^-14^ | 3.578x10^-2^ | 1.534x10^-2^ - 8.341x10^-2^ |
| rs376062400 | 12239 | C>T | *MT-TS2* | 2.849x10^-10^ | 9.782x10^-3^ | 2.322x10^-3^ - 4.12x10^-2^ |
| rs878853101 | 14022 | A>G | *MT-ND5* | 3.093x10^-9^ | 1.288x10^-2^ | 3.052x10^-3^ - 5.431x10^-2^ |
| rs28357678 | 14668 | C>T | *MT-ND6* | 1.096x10^-17^ | 7.356x10^-2^ | 4.048x10^-2^ - 1.337x10^-1^ |
| rs1603225763 | 16247 | A>G | *MT-HV1* | 4.746x10^-10^ | 2.237x10^-2^ | 6.765x10^-3^ - 7.398x10^-2^ |
