## Supplementary material for "Mitochondrial genome variants associated with Amyotrophic Lateral Sclerosis and their haplogroup distribution": Table S10-Heteroplasmy analysis.docx

**Supplementary Table S8.** Heteroplasmy analysis summary (1,978 samples). Heteroplasmy analysis conducted on 1,978 mitogenomes (1,965 cases and 13 controls). Of all evaluated sites, 26.7% were heteroplasmic, while 73.3% are homoplasmic sites.

| **Variant** | | | | | **Variant type** | |
| --- | --- | --- | --- | --- | --- | --- |
|  | A | C | G | T | heteroplasmic | homoplasmic |
| Frequency | 12011 | 16925 | 23159 | 11678 | 17059 | 46714 |
| Proportion | 0.189 | 0.265 | 0.363 | 0.183 | 0.267 | 0.733 |
