## Supplementary material for "Mitochondrial genome variants associated with Amyotrophic Lateral Sclerosis and their haplogroup distribution": Table S11-Functional annotation of non-synonymous substitutions.docx

**Supplementary Table S9.** Functional annotation of non-synonymous mitochondrial substitutions associated with ALS. Description of the columns and thresholds obtained are all described in https://mitimpact.css-mendel.it/result_legend.

| **Position** | **Change** | **MitImpact id** | **Gene symbol** | **Respiratory Chain complex** | **Ensembl gene id** | **Ensembl protein id** | **Ensembl transcript id** |
| --- | --- | --- | --- | --- | --- | --- | --- |
| chrMT:4824 | A>G | MI.13512 | *MT-ND2* | I | ENSG00000198763 | ENSP00000355046 | ENST00000361453 |
| chrMT:5178 | C>A | MI.14253 | *MT-ND2* | I | ENSG00000198763 | ENSP00000355046 | ENST00000361453 |
| chrMT:8414 | C>T | MI.1547 | *MT-ATP8* | V | ENSG00000228253 | ENSP00000355265 | ENST00000361851 |
| chrMT:8684 | C>T | MI.337 | *MT-ATP6* | V | ENSG00000198899 | ENSP00000354632 | ENST00000361899 |
| chrMT:8794 | C>T | MI.569 | *MT-ATP6* | V | ENSG00000198899 | ENSP00000354632 | ENST00000361899 |
| chrMT:13105 | A>G | MI.20880 | *MT-ND5* | I | ENSG00000198786 | ENSP00000354813 | ENST00000361567 |
| chrMT:13928 | G>C | MI.22649 | *MT-ND5* | I | ENSG00000198786 | ENSP00000354813 | ENST00000361567 |
| chrMT:14318 | T>C | MI.23447 | *MT-ND6* | I | ENSG00000198695 | ENSP00000354665 | ENST00000361681 |

**Supplementary Table S7 (Continued)**.

| **Position** | **Change** | **Ncbi gene id** | **Gene position** | **AA pos** | **AA ref** | **AA alt** | **Codon substitution** | **PhyloP 100V** | **PhastCons 100V** |
| --- | --- | --- | --- | --- | --- | --- | --- | --- | --- |
| chrMT:4824 | A>G | 4536 | 355 | 119 | T | A | Acc/Gcc | -14.7 | 0 |
| chrMT:5178 | C>A | 4536 | 709 | 237 | L | M | Cta/Ata | 0.03 | 0 |
| chrMT:8414 | C>T | 4509 | 49 | 17 | L | F | Ctc/Ttc | -2.42 | 0 |
| chrMT:8684 | C>T | 4508 | 158 | 53 | T | I | aCc/aTc | 0.13 | 0 |
| chrMT:8794 | C>T | 4508 | 268 | 90 | H | Y | Cac/Tac | -20 | 0 |
| chrMT:13105 | A>G | 4540 | 769 | 257 | I | V | Atc/Gtc | -8.76 | 0 |
| chrMT:13928 | G>C | 4540 | 1592 | 531 | S | T | aGc/aCc | -4.19 | 0 |
| chrMT:14318 | T>C | 4541 | 356 | 119 | N | S | aAt/aGt | -0.29 | 0 |

**Supplementary Table S7 (Continued)**.

| **Position** | **Change** | **PolyPhen2** | **PolyPhen2 score** | **SIFT** | **SIFT score** | **FatHmmW** | **FatHmmW score** | **FatHmm** | **FatHmm score** |
| --- | --- | --- | --- | --- | --- | --- | --- | --- | --- |
| chrMT:4824 | A>G | benign | 0.36 | neutral | 0.42 | neutral | 4.64 | neutral | -0.37 |
| chrMT:5178 | C>A | possibly damaging | 0.9 | neutral | 0.25 | neutral | 4.67 | neutral | -1.39 |
| chrMT:8414 | C>T | probably damaging | 0.99 | neutral | 0.31 | neutral | 1.71 | neutral | -2.75 |
| chrMT:8684 | C>T | benign | 0.01 | neutral | 0.4 | neutral | 4.53 | neutral | 1.16 |
| chrMT:8794 | C>T | benign | 0 | neutral | 1 | neutral | 4.68 | neutral | 3.37 |
| chrMT:13105 | A>G | benign | 0.01 | neutral | 0.52 | neutral | 4.65 | neutral | 1.31 |
| chrMT:13928 | G>C | benign | 0.01 | neutral | 0.4 | neutral | 0.96 | neutral | -0.45 |
| chrMT:14318 | T>C | benign | 0.02 | neutral | 0.44 | neutral | 2.17 | deleterious | -3.41 |

**Supplementary Table S7 (Continued)**.

| **Position** | **Change** | **PROVEAN** | **PROVEAN score** | **MutationAssessor** | **MutationAssessor score** | **EFIN SP score** | **EFIN SP** | **EFIN HD score** | **EFIN HD** |
| --- | --- | --- | --- | --- | --- | --- | --- | --- | --- |
| chrMT:4824 | A>G | deleterious | -4.25 | low impact | 0.86 | 0.98 | neutral | 0.83 | neutral |
| chrMT:5178 | C>A | neutral | 0.39 | neutral impact | 0.14 | 0.99 | neutral | 0.83 | neutral |
| chrMT:8414 | C>T | neutral | -1.98 | low impact | 1.05 | 1 | neutral | 0.79 | neutral |
| chrMT:8684 | C>T | neutral | -0.23 | neutral impact | -1.06 | 0.97 | neutral | 0.97 | neutral |
| chrMT:8794 | C>T | neutral | 0.24 | neutral impact | -2.38 | 0.96 | neutral | 0.89 | neutral |
| chrMT:13105 | A>G | neutral | 0.02 | neutral impact | -0.72 | 0.94 | neutral | 0.96 | neutral |
| chrMT:13928 | G>C | neutral | 1.36 | neutral impact | -0.7 | 0.92 | neutral | 0.99 | neutral |
| chrMT:14318 | T>C | neutral | -1.98 | neutral impact | 0.66 | 0.93 | neutral | 0.96 | neutral |

**Supplementary Table S7 (Continued)**.

| **Position** | **Change** | **CADD score** | **CADD phred score** | **CADD** | **VEST**  ***p-*value** | **VEST** | **VEST FDR** | **PANTHER score** | **PANTHER** | **PhD.SNP score** | **PhD.SNP** |
| --- | --- | --- | --- | --- | --- | --- | --- | --- | --- | --- | --- |
| chrMT:4824 | A>G | 2.31 | 18.24 | deleterious | 0.21 | neutral | 0.45 | 0.55 | disease | 0.3 | neutral |
| chrMT:5178 | C>A | 2.36 | 18.59 | deleterious | 0.32 | neutral | 0.5 | 0.64 | disease | 0.1 | neutral |
| chrMT:8414 | C>T | 2.42 | 18.98 | deleterious | 0.51005936 | neutral | 0.85 | 0.27 | neutral | 0.15 | neutral |
| chrMT:8684 | C>T | 0.47 | 7.17 | neutral | 0.28 | neutral | 0.65 | 0.29 | neutral | 0.23 | neutral |
| chrMT:8794 | C>T | -0.94 | 0.02 | neutral | 0.7 | neutral | 0.75 | 0.43 | neutral | 0.06 | neutral |
| chrMT:13105 | A>G | -0.58 | 0.14 | neutral | 0.71 | neutral | 0.75 | 0.36 | neutral | 0.11 | neutral |
| chrMT:13928 | G>C | -0.98 | 0.02 | neutral | 0.32 | neutral | 0.5 | 0.28 | neutral | 0.09 | neutral |
| chrMT:14318 | T>C | -0.55 | 0.17 | neutral | 0.48 | neutral | 0.55 | 0.1 | neutral | 0.47 | neutral |

**Supplementary Table S7 (Continued)**.

| **Position** | **Change** | **SNAP score** | **SNAP** | **MutationTaster** | **MutationTaster score** | **Mitoclass1** | **SNPDryad score** | **SNPDryad** | **Meta.SNP score** | **Meta.SNP** | **Meta.SNP RI** |
| --- | --- | --- | --- | --- | --- | --- | --- | --- | --- | --- | --- |
| chrMT:4824 | A>G | 0.49 | neutral | polymorphism | 1 | damaging | 0.84 | neutral | 0.58 | disease | 2 |
| chrMT:5178 | C>A | 0.4 | neutral | polymorphism | 1 | neutral | 0.43 | neutral | 0.43 | neutral | 1 |
| chrMT:8414 | C>T | 0.37 | neutral | polymorphism | 1 | neutral | 0.12 | neutral | 0.06 | neutral | 9 |
| chrMT:8684 | C>T | 0.43 | neutral | polymorphism | 1 | neutral | 0.06 | neutral | 0.41 | neutral | 2 |
| chrMT:8794 | C>T | 0.52 | disease | polymorphism | 1 | neutral | 0.04 | neutral | 0.21 | neutral | 6 |
| chrMT:13105 | A>G | 0.37 | neutral | polymorphism | 1 | neutral | 0 | neutral | 0.29 | neutral | 4 |
| chrMT:13928 | G>C | 0.13 | neutral | polymorphism | 1 | neutral | 0 | neutral | 0.26 | neutral | 5 |
| chrMT:14318 | T>C | 0.42 | neutral | . | . | neutral | 0.1 | neutral | 0.44 | neutral | 1 |

**Supplementary Table S7 (Continued)**.

| **Position** | **Change** | **CAROL score** | **CAROL** | **Condel score** | **Condel** | **COVEC WMV score** | **COVEC WMV** | **MtoolBox DS** | **MtoolBox** | **APOGEE score** | **APOGEE** |
| --- | --- | --- | --- | --- | --- | --- | --- | --- | --- | --- | --- |
| chrMT:4824 | A>G | 0.51 | neutral | 0.53 | deleterious | -6 | neutral | 0.44 | deleterious | 0.4 | neutral |
| chrMT:5178 | C>A | 0.92 | neutral | 0.18 | neutral | -3 | neutral | 0.63 | deleterious | 0.68 | pathogenic |
| chrMT:8414 | C>T | 0.99 | deleterious | 0.16 | neutral | -2 | neutral | 0.71 | deleterious | 0.58 | pathogenic |
| chrMT:8684 | C>T | 0.59 | neutral | 0.7 | deleterious | -6 | neutral | 0.11 | neutral | 0.32 | neutral |
| chrMT:8794 | C>T | 0 | neutral | 1 | deleterious | -6 | neutral | 0.13 | neutral | 0.44 | neutral |
| chrMT:13105 | A>G | 0.47 | neutral | 0.76 | deleterious | -6 | neutral | 0.1 | neutral | 0.32 | neutral |
| chrMT:13928 | G>C | 0.59 | neutral | 0.7 | deleterious | -6 | neutral | 0.32 | neutral | 0.46 | neutral |
| chrMT:14318 | T>C | 0.54 | neutral | 0.71 | deleterious | -6 | neutral | 0.12 | neutral | 0.41 | neutral |

**Supplementary Table S7 (Continued)**.

| **Position** | **Change** | **DEOGEN2 score** | **DEOGEN2** | **PolyPhen2 transf score** | **PolyPhen2 transf** | **SIFT transf score** | **SIFT transf** | **MutationAssessor transf score** | **MutationAssessor transf** |
| --- | --- | --- | --- | --- | --- | --- | --- | --- | --- |
| chrMT:4824 | A>G | 0.06 | neutral | -0.57 | medium impact | 0.13 | medium impact | -0.42 | medium impact |
| chrMT:5178 | C>A | 0 | neutral | -1.67 | low impact | -0.06 | medium impact | -1.03 | low impact |
| chrMT:8414 | C>T | 0.03 | neutral | -2.65 | low impact | 0.1 | medium impact | -0.2 | medium impact |
| chrMT:8684 | C>T | 0.01 | neutral | 1.14 | medium impact | 0.19 | medium impact | -2.01 | low impact |
| chrMT:8794 | C>T | 0.03 | neutral | 2.09 | high impact | 1.98 | high impact | -3.14 | low impact |
| chrMT:13105 | A>G | 0.03 | neutral | 1.15 | medium impact | 0.25 | medium impact | -1.86 | low impact |
| chrMT:13928 | G>C | 0 | neutral | 1.15 | medium impact | 0.14 | medium impact | -1.84 | low impact |
| chrMT:14318 | T>C | 0.31 | neutral | 0.75 | medium impact | 0.15 | medium impact | -0.59 | medium impact |

**Supplementary Table S7 (Continued)**.

| **Position** | **Change** | **CHASM *p-*value** | **CHASM FDR** | **CHASM** | **dbSNP 153 id** | **ClinVar March2020 ClinSig** | **ClinVar March2020 ClnDBN** | **ClinVar March2020 ClnAllele id** |
| --- | --- | --- | --- | --- | --- | --- | --- | --- |
| chrMT:4824 | A>G | 0.34 | 0.8 | neutral | rs1556422903 | benign | Leigh syndrome | 681046 |
| chrMT:5178 | C>A | 0.51 | 0.8 | neutral | rs28357984 | benign | Leigh syndrome | 681087 |
| chrMT:8414 | C>T | 0.65 | 0.85 | neutral | rs28358884 | benign | Leigh syndrome | 681386 |
| chrMT:8684 | C>T | 0.65 | 0.9 | neutral | rs201336180 | benign | Leigh syndrome | 681487 |
| chrMT:8794 | C>T | 0.39 | 0.9 | neutral | rs2298007 | benign | Leigh syndrome | 681523 |
| chrMT:13105 | A>G | 0.58 | 0.8 | neutral | rs2853501 | benign | Leigh syndrome | 680409 |
| chrMT:13928 | G>C | 0.7 | 0.85 | neutral | rs28359184 | benign | Leigh syndrome | 680526 |
| chrMT:14318 | T>C | 0.36 | 0.8 | neutral | rs28357675 | benign | Leigh syndrome | 680599 |

**Supplementary Table S7 (Continued)**.

| **Position** | **Change** | **COSMIC 90 id** | **MITOMAP allele** | **MITOMAP phenotype** | **MITOMAP homoplasmy** | **MITOMAP heteroplasmy** | **MITOMAP status** | **MITOMAP NRef** |
| --- | --- | --- | --- | --- | --- | --- | --- | --- |
| chrMT:4824 | A>G | . | . | . | . | . | . | . |
| chrMT:5178 | C>A | . | C5178A | Longevity / Extraversion / diabetes / AMS protection / blood iron metabolism / correlation with myocardial infarction / atherosclerosis | + | - | Reported | 23 |
| chrMT:8414 | C>T | . | C8414T | Increased risk of T2DM in haplogroup D4 / Longevity | + | - | Reported | 1 |
| chrMT:8684 | C>T | . | . | . | . | . | . | . |
| chrMT:8794 | C>T | COSM488775 | C8794T | Exercise Endurance / Coronary Atherosclerosis risk | + | - | Reported | 2 |
| chrMT:13105 | A>G | COSM6716789 | . | . | . | . | . | . |
| chrMT:13928 | G>C | . | . | . | . | . | . | . |
| chrMT:14318 | T>C | . | . | . | . | . | . | . |
