## Supplementary material for "Mitochondrial genome variants associated with Amyotrophic Lateral Sclerosis and their haplogroup distribution": Table S12-SNVs associations with other diseases.docx

**Supplementary Table S10.** Mitochondrial variants associated with ALS and their previously reported associations with other diseases. Only variants with *p*-value < 10^-7^ are shown (Table 1 and Table 2). *MT* = mitochondrial, *HV1* = hyper variable region 1, *HV2* = hyper variable region 2, *HV3* = hyper variable region 3, *ND* = NADH ubiquinone oxidoreductases, *RNR1* = 12S ribosomal RNA, *RNR2* = 16S ribosomal RNA, *ATP6* = mitochondrially encoded ATP synthase membrane subunit 6, *ATP8* = mitochondrially encoded ATP synthase membrane subunit 8, *CYB* = Cytochrome b, LVNC = left ventricular noncompaction, AMS = acute mountain sickness, T2DM = type 2 diabetes and MDD = major depressive disorder. Variants were annotated using the MITOMAP (<https://www.mitomap.org/MITOMAP>), with dbSNP ID numbers confirmed in dbSNP build 152 ([www.ncbi.nlm.nih.gov/snp/](http://www.ncbi.nlm.nih.gov/snp/)).

| **dbSNP ID** | **Position** | **Change** | **Gene** | **Disease Association** | **PubMed IDs** |
| --- | --- | --- | --- | --- | --- |
| rs3901846 | 499 | G>A | *MT-HV3* | Endometriosis / possible protective factor for high altitude sickness | 29124462, 23096691 |
| rs56489998 | 663 | A>G | *MT-RNR1* | Coronary atherosclerosis risk | 21099167 |
| rs28358579 | 2352 | T>C | *MT-RNR2* | Possibly LVNC associated | 27498855, 20211276, 27217714 |
| rs28358587 | 3552 | T>A | *MT-ND1* | Resistance to high altitude pulmonary edema (HAPE) | 23350576 |
| rs200763872 | 4883 | C>T | *MT-ND2* | Glaucoma | 27217714, 24448266 |
| rs28357984 | 5178 | C>A | *MT-ND2* | Longevity / Extraversion / diabetes / AMS protection / blood iron metabolism / correlation with myocardial infarction / atherosclerosis | 9449878, 10996007, 11735027, 12391595, 12375058, 12384792, 12782420, 15126279, 15211636, 15262184, 16271520, 19667492, 20555337, 20306229, 21319252, 21385625, 18468491, 11573146, 28951770, 29670672, 25834827, 29987491, 31488191 |
| rs28358884 | 8414 | C>T | *MT-ATP8* | Increased risk of T2DM and high-altitude polycythemia (HAPC) in haplogroup D4 / Longevity | 18468491, 33420243, 24498190 |
| rs2298007 | 8794 | C>T | *MT-ATP6* | Exercise Endurance / Coronary Atherosclerosis risk | 15126279, 21099167 |
| rs878969753 | 10142 | C>T | *MT-ND3* | Recurrent pregnancy loss | 28696810 |
| rs28357678 | 14668 | C>T | *MT-ND6* | Depressive Disorder associated | 19290059 |
| rs28357680 | 14783 | T>C | *MT-CYB* | Possible role in high altitude sickness | 33420243 |
| rs193302985 | 15043 | G>A | *MT-CYB* | MDD associated / possible role in high altitude sickness | 19290059, 16773565, 33420243 |
| rs193302991 | 15301 | G>A | *MT-CYB* | Possible role in high altitude sickness | 33420243 |
| rs35134837 | 16217 | T>C | *MT-HV1* | Endometriosis | 29124462 |
