## Supplementary material for "Mitochondrial genome variants associated with Amyotrophic Lateral Sclerosis and their haplogroup distribution": The NYGC ALS Consortium.docx

1. Hemali Phatnani, Center for Genomics of Neurodegenerative Disease (CGND), New York Genome Center, New York, NY
2. Justin Kwan, Department of Neurology, Lewis Katz School of Medicine, Temple University, Philadelphia, PA
3. Dhruv Sareen, Cedars-Sinai Department of Biomedical Sciences, Board of Governors Regenerative Medicine Institute and Brain Program, Cedars-Sinai Medical Center, and Department of Medicine, University of California, Los Angeles, CA
4. James R. Broach, Department of Biochemistry and Molecular Biology, Penn State Institute for Personalized Medicine, The Pennsylvania State University, Hershey, PA
5. Zachary Simmons, Department of Neurology, The Pennsylvania State University, Hershey, PA
6. Ximena Arcila-Londono, Department of Neurology, Henry Ford Health System, Detroit, MI
7. Edward B. Lee, MD, Department of Pathology and Laboratory Medicine, Perelman School of Medicine, University of Pennsylvania, Philadelphia, PA
8. Vivianna M. Van Deerlin, Department of Pathology and Laboratory Medicine, Perelman School of Medicine, University of Pennsylvania, Philadelphia, PA
9. Neil A. Shneider, MD, Department of Neurology, Center for Motor Neuron Biology and Disease, Institute for Genomic Medicine, Columbia University, New York, NY
10. Ernest Fraenkel, Department of Biological Engineering, Massachusetts Institute of Technology, Cambridge, MA
11. Lyle W. Ostrow, MD, Department of Neurology, Johns Hopkins School of Medicine, Baltimore, MD
12. Frank Baas, Department of Neurogenetics, Academic Medical Centre, Amsterdam and Leiden University Medical Center, Leiden, The Netherlands
13. Noah Zaitlen, Department of Medicine, Lung Biology Center, University of California, San Francisco, CA
14. James D. Berry, ALS Multidisciplinary Clinic, Neuromuscular Division, Department of Neurology, Harvard Medical School, and Neurological Clinical Research Institute, Massachusetts General Hospital, Boston, MA
15. Andrea Malaspina, Centre for Neuroscience and Trauma, Blizard Institute, Barts and The London School of Medicine and Dentistry, Queen Mary University of London, London, and Department of Neurology, Basildon University Hospital, Basildon, United Kingdom
16. Pietro Fratta, MD, Institute of Neurology, National Hospital for Neurology and Neurosurgery, University College London, London, United Kingdom
17. Gregory A. Cox, The Jackson Laboratory, Bar Harbor, ME
18. Leslie M. Thompson, Department of Psychiatry & Human Behavior, Department of Biological Chemistry, School of Medicine, and Department of Neurobiology and Behavior, School of Biological Sciences, University California, Irvine, CA
19. Steve Finkbeiner, Taube/Koret Center for Neurodegenerative Disease Research, Roddenberry Center for Stem Cell Biology and Medicine, Gladstone Institute
20. Efthimios Dardiotis, Department of Neurology & Sensory Organs, University of Thessaly, Thessaly, Greece
21. Timothy M. Miller, MD, Department of Neurology, Washington University in St. Louis, St. Louis, MO
22. Siddharthan Chandran, Centre for Clinical Brain Sciences, Anne Rowling Regenerative Neurology Clinic, Euan MacDonald Centre for Motor Neurone Disease Research, University of Edinburgh, Edinburgh, United Kingdom
23. Suvankar Pal, Centre for Clinical Brain Sciences, Anne Rowling Regenerative Neurology Clinic, Euan MacDonald Centre for Motor Neurone Disease Research, University of Edinburgh, Edinburgh, United Kingdom
24. Eran Hornstein, Department of Molecular Genetics, Weizmann Institute of Science, Rehovot, Israel
25. Daniel J. MacGowan, Department of Neurology, Icahn School of Medicine at Mount Sinai, New York, NY
26. Terry Heiman-Patterson, Center for Neurodegenerative Disorders, Department of Neurology, the Lewis Katz School of Medicine, Temple University, Philadelphia, PA
27. Molly G. Hammell, Cold Spring Harbor Laboratory, Cold Spring Harbor, NY
28. Nikolaos. A. Patsopoulos, Computer Science and Systems Biology Program, Ann Romney Center for Neurological Diseases, Department of Neurology and Division of Genetics in Department of Medicine, Brigham and Women’s Hospital, Boston, MA, Harvard Medical School, Boston, MA, and Program in Medical and Population Genetics, Broad Institute, Cambridge, MA
29. Oleg Butovsky, Ann Romney Center for Neurologic Diseases, Brigham and Women's Hospital, Harvard Medical School, Boston, MA
30. Joshua Dubnau, Department of Anesthesiology, Stony Brook University, Stony Brook, NY
31. Avindra Nath, Section of Infections of the Nervous System, National Institute of Neurological Disorders and Stroke, NIH, Bethesda, MD
32. Robert Bowser, Department of Neurology, Barrow Neurological Institute, St. Joseph's Hospital and Medical Center, Department of Neurobiology, Barrow Neurological Institute, St. Joseph's Hospital and Medical Center, Phoenix, AZ
33. Matt Harms, Department of Neurology, Division of Neuromuscular Medicine, Columbia University, New York, NY
34. Eleonora Aronica, Department of Neuropathology, Academic Medical Center, University of Amsterdam, Amsterdam, The Netherlands
35. Mary Poss, DVM, Department of Biology and Veterinary and Biomedical Sciences, The Pennsylvania State University, University Park, PA
36. Jennifer Phillips-Cremins, New York Stem Cell Foundation, Department of Bioengineering, School of Engineering and Applied Sciences, University of Pennsylvania, Philadelphia, PA
37. John Crary, MD, Department of Pathology, Fishberg Department of Neuroscience, Friedman Brain Institute, Ronald M. Loeb Center for Alzheimer's Disease, Icahn School of Medicine at Mount Sinai, New York, NY
38. Nazem Atassi, Department of Neurology, Harvard Medical School, Neurological Clinical Research Institute, Massachusetts General Hospital, Boston, MA
39. Dale J. Lange, Department of Neurology, Hospital for Special Surgery and Weill Cornell Medical Center, New York, NY
40. Darius J. Adams, Medical Genetics, Atlantic Health System, Morristown Medical Center, Morristown, NJ, and Overlook Medical Center, Summit, NJ
41. Leonidas Stefanis, Center of Clinical Research, Experimental Surgery and Translational Research, Biomedical Research Foundation of the Academy of Athens (BRFAA), 4 Soranou Efesiou Street, 11527, Athens, Greece; 1st Department of Neurology, Eginition Hospital, Medical School, National and Kapodistrian University of Athens, Athens, Greece
42. Marc Gotkine, Neuromuscular/EMG service and ALS/Motor Neuron Disease Clinic, Hebrew University-Hadassah Medical Center, Jerusalem, Israel
43. Robert H. Baloh, Board of Governors Regenerative Medicine Institute, Los Angeles, CA; Department of Neurology, Cedars-Sinai Medical Center, Los Angeles, CA
44. Suma Babu, MBBS, Neurological Clinical Research Institute, Massachusetts General Hospital, Boston, MA
45. Towfique Raj, Departments of Neuroscience, and Genetics and Genomic Sciences, Ronald M. Loeb Center for Alzheimer's disease, Icahn School of Medicine at Mount Sinai, New York, NY
46. Sabrina Paganoni, Harvard Medical School, Department of Physical Medicine & Rehabilitation, Spaulding Rehabilitation Hospital, Boston, MA
47. Ophir Shalem, Center for Cellular and Molecular Therapeutics, Children's Hospital of Philadelphia, Philadelphia, PA; Department of Genetics, Perelman School of Medicine, University of Pennsylvania, Philadelphia, PA
48. Colin Smith, Centre for Clinical Brain Sciences, University of Edinburgh, Edinburgh, UK; Euan MacDonald Centre for Motor Neurone Disease Research, University of Edinburgh, Edinburgh, UK
49. Bin Zhang, Department of Genetics and Genomic Sciences, Icahn Institute of Data Science and Genomic Technology, Icahn School of Medicine at Mount Sinai, New York, NY
50. University of Maryland Brain and Tissue Bank and NIH NeuroBioBank
51. Brent Harris, Department of Neuropathology, Georgetown Brain Bank, Georgetown Lombardi Comprehensive Cancer Center, Georgetown University Medical Center, Washington DC
52. Iris Broce, Neuroradiology Section, Department of Radiology and Biomedical Imaging, University of California, San Francisco, San Francisco, CA
53. Vivian Drory, Neuromuscular Diseases Unit, Department of Neurology, Tel Aviv Sourasky Medical Center, Sackler Faculty of Medicine, Tel-Aviv University, Tel-Aviv, Israel
54. John Ravits, Department of Neuroscience, University of California San Diego, La Jolla, CA
55. Corey McMillan, Department of Neurology, University of Pennsylvania Perelman School of Medicine, Philadelphia, PA
56. Vilas Menon, Department of Neurology, Columbia University Medical Center, New York, NY
57. Lani Wu, Department of Pharmaceutical Chemistry, University of California San Francisco, San Francisco, CA
58. Steven Altschuler, Department of Pharmaceutical Chemistry, University of California San Francisco, San Francisco, CA.
